# Peanut oral immunotherapy differentially suppresses clonally distinct subsets of T helper cells

**DOI:** 10.1101/2021.07.29.21261049

**Authors:** Brinda Monian, Ang A. Tu, Bert Ruiter, Duncan M. Morgan, Patrick M. Petrossian, Neal P. Smith, Todd M. Gierahn, Julia H. Ginder, Wayne G. Shreffler, J. Christopher Love

**Author notes:** These authors contributed equally (Brinda Monian, Ang A. Tu). These authors contributed equally (Wayne G. Shreffler, J. Christopher Love).

## Abstract

Food allergy affects an estimated 8% of children in the US, with increasing severity and global prevalence^1^. Oral immunotherapy (OIT) is a recently approved treatment with outcomes ranging from sustained tolerance to food allergen to no apparent benefit^2,3^. The immunological underpinnings that influence clinical outcomes of OIT still remain largely unresolved. Using single-cell RNA sequencing and paired TCRα/β sequencing, we assessed the transcriptomes of CD154+ and CD137+ peanut-reactive T helper cells from 12 peanut-allergic patients longitudinally throughout OIT. We observed expanded populations of cells expressing Th1, Th2, and Th17 signatures that further separated into six clonally distinct subsets, including a Tfh1-like, a Tfh2-like, a Th2A-like, and a Th2reg-like subset. Four of these subsets demonstrated convergence of TCR sequences, suggesting antigen-driven T cell fate. Although we observed suppression during OIT of Th2 and Th1 gene signatures within effector clonotypes, Tfh clonotypes were unaffected. We also did not observe significant clonal deletion or induction among the antigen-reactive T cells characterized. Positive outcomes were associated with larger decrease of Th2 signatures in Th2A-like cells, while treatment failure was associated with high baseline inflammatory gene signatures that were unmodulated by OIT. These signatures, including expression of *OX40*, *OX40L, STAT1*, and *GPR15*, were most clearly present in Th1 and Th17 clonotypes, but were also more broadly detected across the CD154+ CD4 population. These results demonstrate that differential clinical response is associated both with pre-existing trait characteristics of the CD4 immune compartment and with susceptibility to modulation by OIT.

**Conflict of Interest Statement:** A.A.T., T.M.G., J.C.L., and the Massachusetts Institute of Technology have filed patents related to the single-cell sequencing methods used in this work. J.C.L. has interests in Sunflower Therapeutics PBC, Pfizer, Honeycomb Biotechnologies, OneCyte Biotechnologies, SQZ Biotechnologies, Alloy Therapeutics, QuantumCyte, Amgen, and Repligen. J.C.L.’s interests are reviewed and managed under Massachusetts Institute of Technology’s policies for potential conflicts of interest. J.C.L. receives sponsored research support at MIT from Amgen, the Bill & Melinda Gates Foundation, Biogen, Pfizer, Roche, Takeda, and Sanofi. The spouse of J.C.L. is an employee of Sunflower Therapeutics PBC. T.M.G. is currently an employee of Honeycomb Biotechnologies, Inc. A.A.T. is currently an employee of Immunitas Therapeutics, Inc. W.G.S. is a consultant of Aimmune Therapeutics.

## Introduction

Food allergy is an immune hypersensitivity condition characterized by high-affinity allergen-specific IgE antibodies and allergen-specific Th2 cells^2,4^. Specific IgE binds to effector cells, such as mast cells and basophils, through FcεRI receptors that are cross-linked upon binding of allergen. The resulting cellular degranulation causes local and systemic release of histamine and other mediators, leading to allergic reactions ranging from mild symptoms, such as hives and abdominal pain, to potentially life-threatening anaphylaxis^5^. Allergen-specific Th2 cells constitute a critical component in this cascade of events. Th2 cells are broadly defined by the expression of the transcription factor GATA3 and the secretion of cytokines IL-4, IL-5, and IL-13, which promote class-switching of B cells to IgE and recruitment of eosinophils as well as other effector cells^6,7^. Recent studies have highlighted subtypes of Th2 cells with specialized functions in the context of allergy, including effector memory (e.g. Th2A, peTh2), and T follicular helper (e.g. Tfh13) phenotypes^7–12^.

Oral immunotherapy (OIT) is currently the only FDA-approved treatment for food allergy intended to prevent anaphylaxis^3^. OIT involves the daily ingestion of escalating doses of allergen. Most patients (80-85%) achieve desensitization (a loss in clinical reactivity with regular consumption of allergen), but only about a third of patients maintain unresponsiveness if treatment is discontinued for even just a few months^13–15^. Studies of the impact of OIT on circulating T cells have consistently found evidence for suppression of Th2 responses, but most of these studies have not correlated T cell responses with heterogenous clinical outcomes^8,16–19^. Similarly, while regulatory T cell (Treg) induction has been observed using *in vitro* expansion of T cells from OIT patients, it has not been consistently shown *ex vivo*^18–25^. Studying allergen-reactive T cell subsets *ex vivo* is challenging due to their low frequencies in peripheral blood and technical limitations to both reliably phenotype these populations and track corresponding clonotypes longitudinally^23,26^. As a result, existing data on T cell responses in the context of OIT have been limited to features comprising a narrow set of genes and proteins or cells specific to a pre-defined subset of allergen epitopes. Comprehensive characterization of allergen-specific CD4+ T cell subsets and their response to immunotherapy over time may not only refine treatment strategies for food allergy, but also enhance our broader understanding of T helper cell phenotypes in atopic disease.

## Results

### Single-cell RNA-Seq enables deep profiling of peanut-reactive T helper cells from OIT patients

To measure the impact of OIT on peanut-reactive T cells, we profiled longitudinal blood samples from 12 patients participating in a clinical trial of peanut OIT (NCT01750879, **Supplementary Table** 1,2). In brief, PBMCs were isolated at four timepoints from each patient: baseline (BL; before therapy), buildup (BU; 13 weeks after start of therapy), maintenance (MN; 12 weeks after the maximum dose was reached), and avoidance (AV; 12 weeks after the end of therapy). Clinical outcomes were evaluated by two oral food challenges and were defined as: tolerance (TO) - passing both food challenges; partial tolerance (PT) - passing the maintenance challenge but failing the avoidance challenge; and treatment failure (TF) - failing the maintenance challenge. Samples from three placebo group (PL) patients were also included (**Figure** 1A; **Methods**).

**Figure 1.**
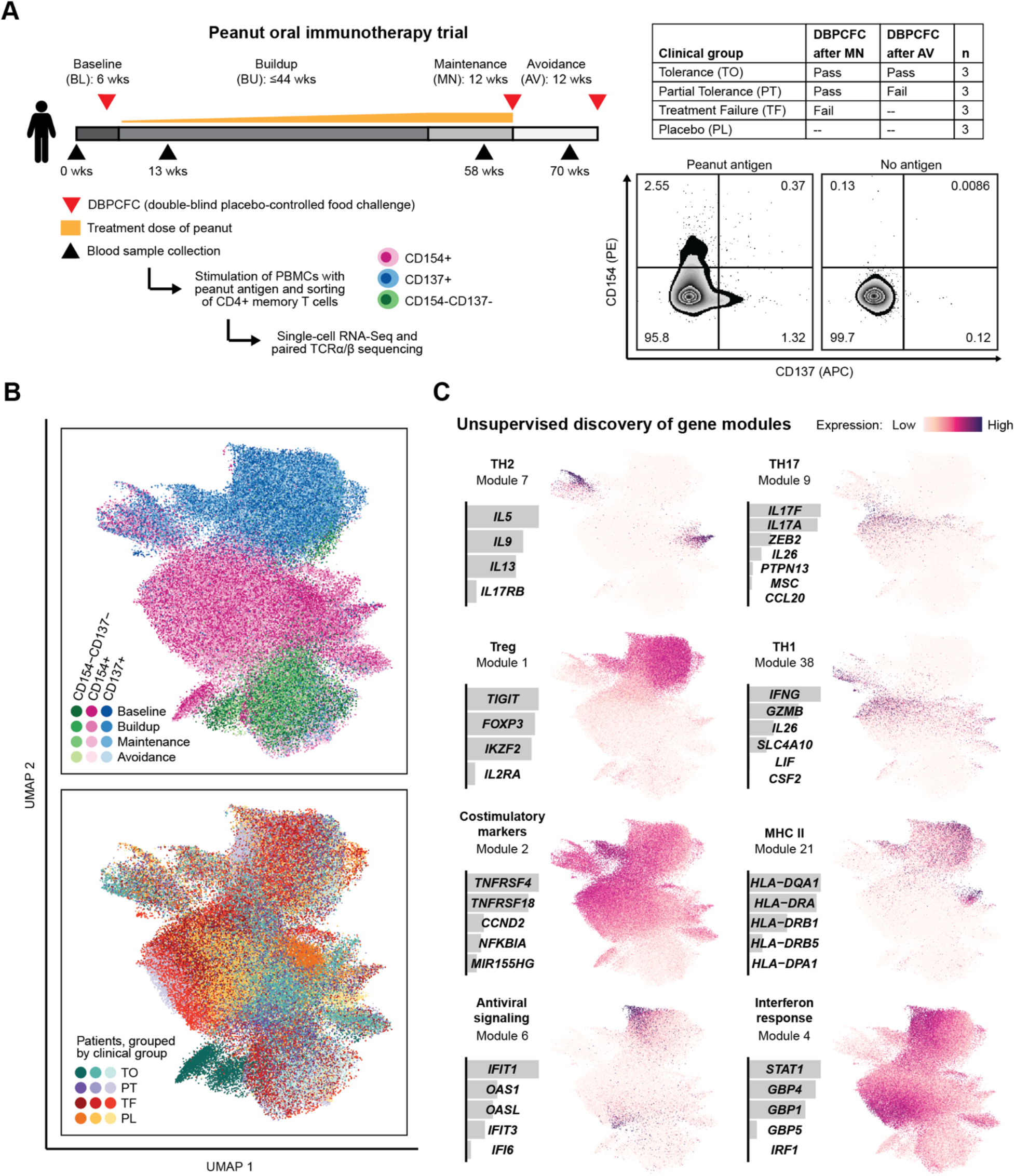
Peanut-reactive T cells from individuals undergoing OIT have diverse transcriptional signatures. **a**, OIT study design, definition of clinical outcomes, and representative flow plots from one patient at baseline. CD3+CD4+CD45RA-memory T cells were sorted by FACS as CD154+CD137+/− (“CD154+”), CD154-CD137+ (“CD137+”), or CD154-CD137-. **b**, Two-dimensional UMAP visualization of all single-cell transcriptomes, colored by sorted subset and time point (top) or by patient and clinical group (bottom). **c**, Selected gene modules discovered using sparse principal components analysis. For each module, a description, the relative weights of each contributing gene, and the module score of all cells overlaid on the UMAP coordinates are shown.

To enrich for allergen-specific T cells and capture their activated profiles, we cultured the PBMCs with whole peanut protein extract for 20 hours to activate CD4+ memory T cells. Peanut-reactive cells were then enriched via FACS using CD154 and CD137 (activation markers for Teff and Treg states, respectively) (**Figure** 1A; **Supplementary Figure** 1A). This approach allowed us to recover a broad set of peanut-specific T cells with limited bias for specific epitopes or HLA types^27^. The relatively short stimulation time was intended to capture *ex vivo* cell states and reflect *in vivo* clonal distributions^28^. CD154-based approaches have been broadly used to identify antigen-reactive CD4+ T cells in various contexts^26,29,30^. In addition, we have previously shown that the frequency of peanut-reactive CD4+ T cells, identified by CD154 expression, is correlated with severity of peanut allergy^11^. Using this method, we observed that OIT significantly decreased the frequency of peanut-reactive CD154+ and CD137+ T cells in the peripheral blood. (**Supplementary Figure** 1B). To further characterize the peanut-reactive memory CD4+ T cells and study how their phenotypes and repertoire are altered during OIT in relation to treatment outcome, we processed the sorted cells for single-cell RNA-Seq via Seq-Well and paired single-cell TCRα/βsequencing^31,32^. In total, we recovered high-quality transcriptomes for 134,129 cells (74,646 CD154+, 41,186 CD137+, and 18,297 CD154-CD137-; **Methods; Supplementary Figure** 2).

Peanut-reactive T cell transcriptomes formed clusters associated most closely with their sorted subsets (**Figure** 1B). CD154+ and CD137+ cells were separated as expected by several differentially expressed genes, including their associated transcripts *CD40LG* and *TNFRSF9*, and other transcripts consistent with their respective effector or regulatory phenotypes (**Extended Data Figure** 1A). We observed patient-specific variation within each cluster that was not a function of library size or mitochondrial content (**Figure** 1B; **Supplementary Figure** 2B), suggesting that it represented inherent biological rather than technical differences. Qualitatively, there was no strong association between transcriptome (as measured by UMAP embedding) and time point or treatment outcome, suggesting that OIT-induced effects might be subtle rather than dominant in the data.

### Sparse PCA delineates canonical and new T-helper cell gene modules

To uncover evidence of OIT-driven variation among peanut-reactive T cells, we developed an unsupervised approach to identify conserved programs of immune-related gene expression. The dataset was filtered to 937 immune and variable genes (**Supplementary Table** 3). Then, co-expressed genes were aggregated into gene modules using sparse principal components analysis (PCA)^33^ to derive a set of 50 gene modules (**Methods**; **Supplementary Figure** 3,4). Several modules corresponded well with the phenotypes of known T cell subsets, such as Th1, Th2, Th17, and regulatory T cells (**Figure** 1C). 43 out of 50 gene modules were present across most or all patients (**Supplementary Figure** 5; **Methods**), suggesting that these represent programs of T cell function or activation that are consistent among individuals.

### Th-related gene modules are associated with expanded T cells

To investigate clonal T cell responses to peanut antigens, we recovered paired TCR sequences from the single-cell transcriptomic sequencing libraries. We identified TCRβ sequences for 60% (+/−17%), TCRα for 55% (+/−15%) and both chains for 36% of cells (+/− 12%) (numbers represent median +/− standard deviation across patients). Coverage was uniform across samples, and the majority of expanded TCRβ sequences were paired with a single TCRα (**Figure** 2A; **Supplementary Figure** 6). Given this relationship, we used TCRβ for all subsequent analyses involving clonotypes. The diversities of CD154+ and CD137+ repertoires were significantly lower than those of the CD154-CD137-cells, indicating that these activation markers enriched for a pool of clonally expanded, peanut-reactive clonotypes (**Figure** 2B, C). In addition, we observed that 55% of expanded clones were detected across multiple time points, but only 1.6% of clonotypes were shared between CD154+ and CD137+ cells, suggesting that these two activated subsets resulted from fundamental differences in lineage, epitope specificity, or both (**Figure** 2D).

**Figure 2.**
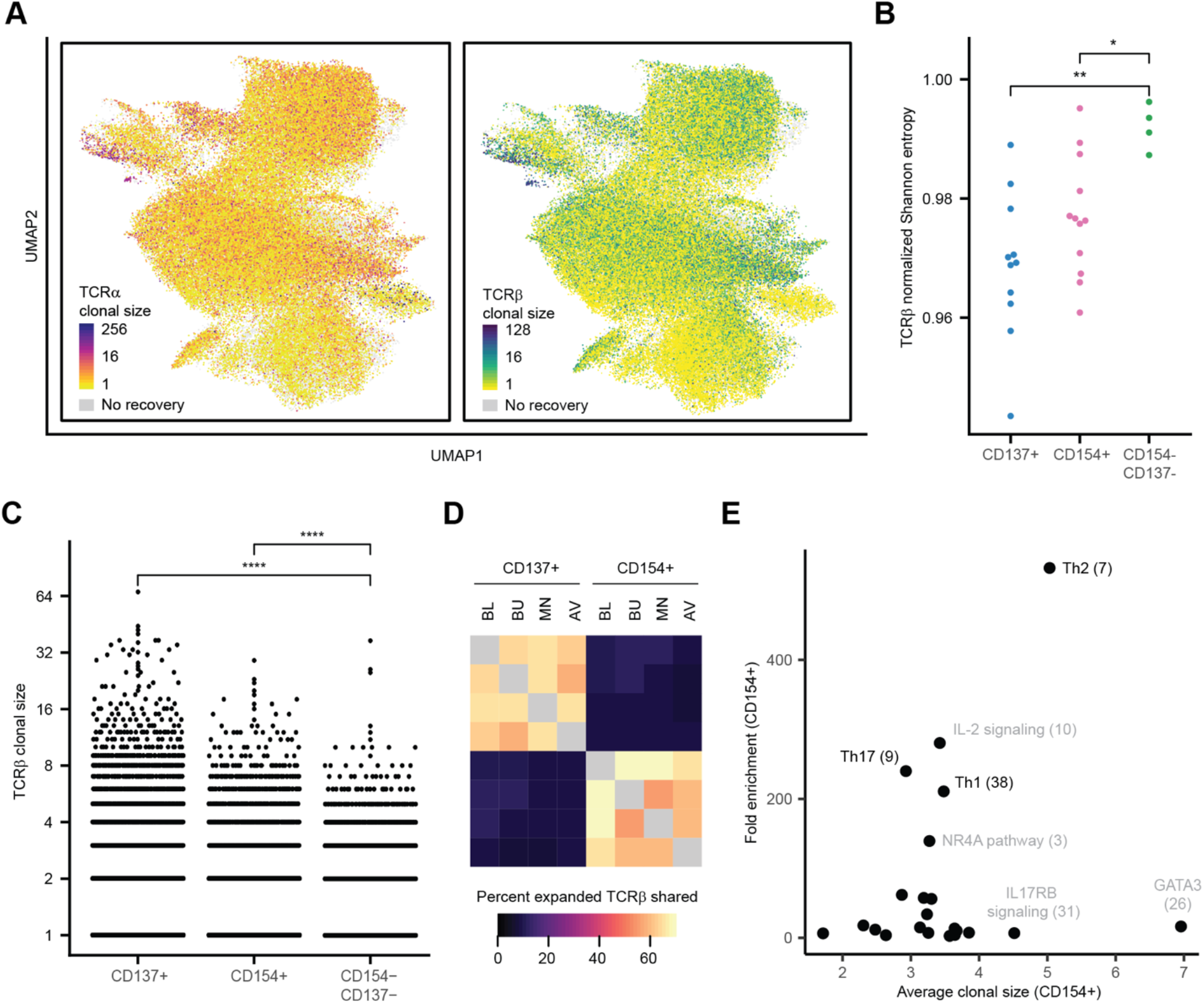
Gene modules for T helper function are associated with clonal expansion and expression in activated cells. **a**, Clonal size of TCRα sequence (left) or TCRβ sequence (right) for all cells with paired TCR recovery, overlaid onto UMAP coordinates. Clonal size is defined as the number of cells sharing a TCR sequence. **b**, Normalized diversity (Shannon index) of TCRβ repertoires of each sorted subset. Each data point represents the repertoire of one patient at all time points. ‘**’ refers to an adjusted p-value of <0.01 by Wilcoxon rank-sum test, and ‘*’ refers to an adjusted p-value of <0.05. **c**, Distribution of TCRβ clonal sizes, within each sorted subset. ‘****’ refers to an adjusted p-value of <0.0001 by Wilcoxon rank-sum test. **d**, Percent of TCRβ sequences shared between time points and sorted subsets. ‘Percent shared’ is defined as the number of unique TCRβ sequences detected in both conditions, divided by the geometric mean of the number of unique TCRβ sequences in each of the two conditions. Sequences from all treatment-group patients were pooled. **e**, Mean clonal size and fold-change in mean module scores (compared to module-expressing CD154-CD137-cells) in CD154+ cells expressing each gene module. Cells were classified as ‘expressing’ each module or not, relative to background expression (**Methods**). Clonal size was calculated with respect to all cells in the dataset.

To determine which, if any, gene modules were associated with clonal T cell expansion, we classified cells as expressing or non-expressing for each module, based on whether the module score was above background expression in CD154-CD137-cells (**Methods**). We then calculated the average TCRβ clonal size for cells expressing each module, as well as the average score of that module in CD154+ cells relative to CD154-CD137-cells. We found that modules representing Th1, Th2, and Th17 functions exhibited strong upregulation in both the CD154+ and CD137+ compartments and were associated with expanded T cell clonotypes (**Figure** 2E; **Extended Data Figure** 1).

### T helper cells comprise six clonally distinct subtypes

Due to their strong enrichment in the CD154+ and CD137+ compartments, we further analyzed the heterogeneity among cells expressing the Th1, Th2, and Th17 modules. Separate clustering of these cells revealed three phenotypically distinct clusters of Th2 cells and two clusters of Th1 cells. We did not observe additional clusters within the Th17 cells (**Figure** 3A). These clusters were detected in all patients (**Supplementary Figure** 7C). Within the Th2 cells, the clusters corresponded to a Tfh2-like (high in costimulatory markers, *CXCR5*, and *PDCD1*), a Th2reg-like (*FOXP3* and *TNFRSF9*), and a Th2A-like population^8,34^ (*GATA3*, *IL17RB*, and *PTGDR2*; **Figure** 3B). The Tfh2-like population resembled a previously-described pathogenic Tfh13 subset, while the Th2A-like population shared markers previously identified in Th2A and peTh2 populations^8–10^ (**Supplementary Figure** 8). Likewise, the Th2reg-like population shared features with previously described deviated Treg cells in food allergy^35^. Among the Th1 cells, the clusters corresponded to a Tfh1-like population and a Th1-conv (conventional) population with canonical Th1 signatures^36^ (**Figure** 3A, B). Both of these clusters expressed high levels of *IFNG* and *GZMB*, and the Tfh1-like cluster exhibited high overlap of genes also expressed in the Tfh2-like population, including *ICOS*, *PDCD1* and *TNFRSF9*.

**Figure 3.**
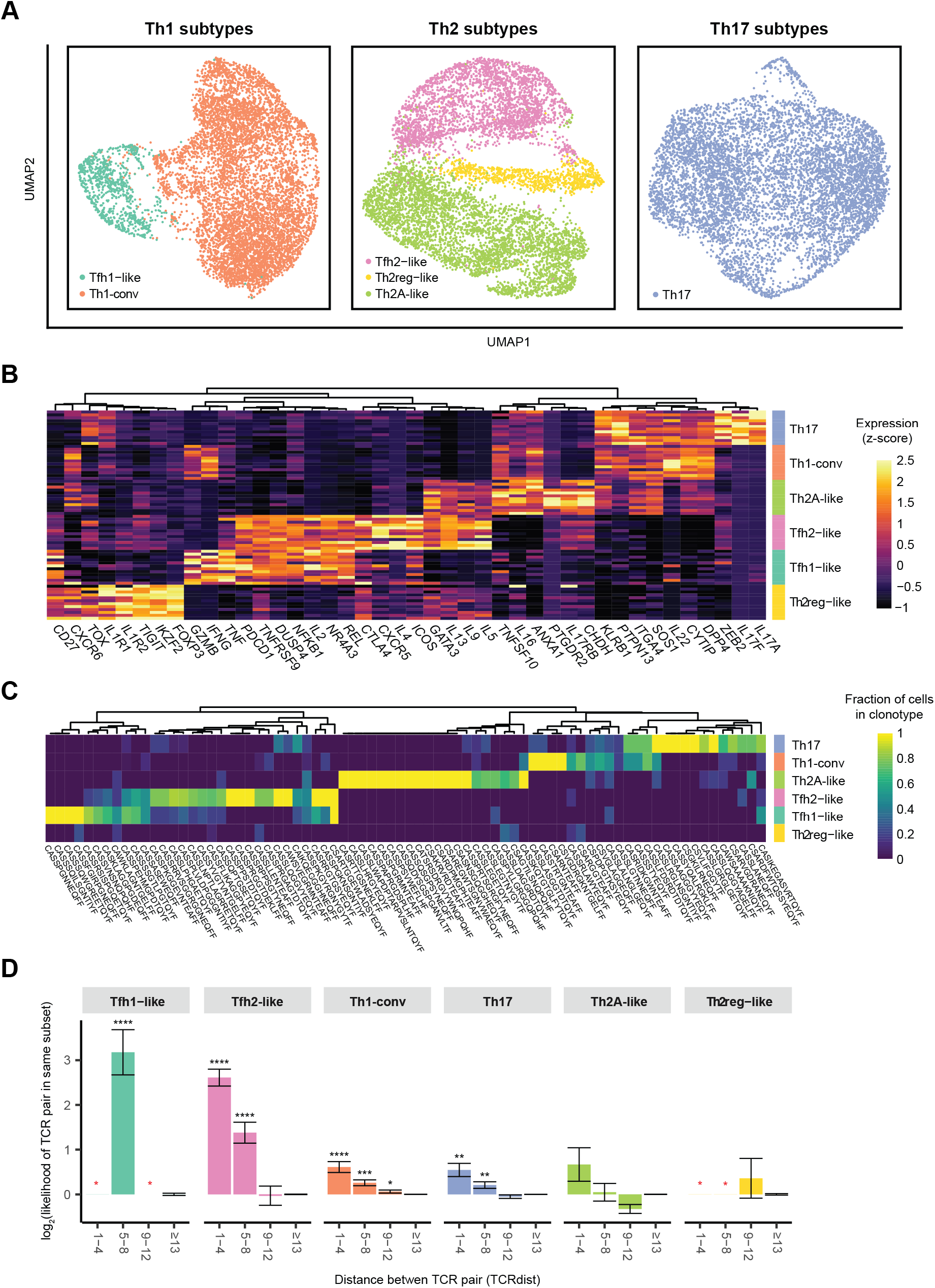
Peanut-reactive T helper subtypes are clonally distinct and exhibit TCR convergence. **a,** UMAP visualizations of Th1-, Th2-, and Th17-scoring cells. Clusters are annotated by their putative identity. **b**, Differentially expressed genes in each T helper subset. Genes were selected using ROC test and manual curation. Each row represents the scaled average gene expression in one patient. **c**, Fraction of TCRβ clonotypes belonging to each subset. Fraction is defined as the number of cells of a TCRβ sequence (column) detected in each subset, divided by the total number of cells within the clonotype. Clonotypes were randomly downsampled to visualize a comparable number from each subset. **d**, TCR distance analysis of TCRβ sequences in the subsets. x-axis represents bins of increasing pairwise TCR distance, calculated using TCRdist. y-axis represents the likelihood for pairs of cells with a given TCR distance to be of the same T helper subset, normalized to the prior probability of any two cells belonging to that subset (see **Methods**). ‘****’ refers to an adjusted p-value of <0.0001 by a two-sided Chi-square proportion test with one degree of freedom, ‘***’ refers to p-value of <0.001, and ‘**’ refers to p-value of <0.01. Data represent combined data from all patients at all timepoints (**a**-**d**).

Analysis of the TCR repertoires of the six T-helper subtypes showed that most clones were primarily associated with a single subtype, indicating that these populations represent distinct clonal lineages (**Figure** 3C). We did, however, observe overlapping clones between the Th1-conv and Th17 states as well as the Tfh1-like and Tfh2-like states, suggesting that cells may transition between these pairs of phenotypic states, or that these states may include shared cellular lineages that differentiated relatively late^37^.

To determine to what extent this association between clonotype and phenotype might be influenced by epitope recognition, we next assessed whether or not TCRs showed evidence of convergence within T helper subtypes using TCRdist, a quantitative metric for similarity between a pair of TCR sequences^38^ (**Methods**). A pair of cells with very similar TCR sequences may share epitope binding properties despite having different ancestries, allowing an assessment of the role of epitope recognition in shaping T cell phenotypes. We found that pairs of cells with highly similar TCRβ sequences (TCRdist < 9) had a significantly enriched likelihood of both cells belonging to the same T helper subtype (p<0.05 by a Chi-square proportion test), with the exception of cells in the Th2A-like and Th2reg-like subtypes (**Figure** 3D). This result indicates a convergence onto common TCR motifs within most subtypes and suggests that factors such as TCR affinity or antigen context during priming (e.g. local tissue environment) may influence the induction of specific T helper phenotypes within an individual^39–41^.

### OIT suppresses Th2 and Th1 signatures in conventional effector, but not Tfh-like, cells

We next assessed the impact of OIT on the TCR repertoire and the identified T helper subtypes. The majority of expanded CD154+ and CD137+ clonotypes were present at three or four of the timepoints, and no timepoint was associated with the depletion or emergence of unique expanded clonotypes or singletons, suggesting that OIT does not induce strong changes in peanut-reactive TCR repertoires (**Extended Data Figure** 2). To investigate the impact of OIT on functional T cell phenotypes, we assessed the mean expression of Th1, Th2, and Th17 modules in all CD154+ cells from each patient and found evidence of suppression at the bulk-level in Th2 and, to a lesser extent, Th1 module scores (adjusted p-values of 0.036 and 0.117, respectively) between the baseline and maintenance timepoints (**Figure** 4A). This was not observed in patients treated with placebo (**Supplementary Figure** 9B).

**Figure 4.**
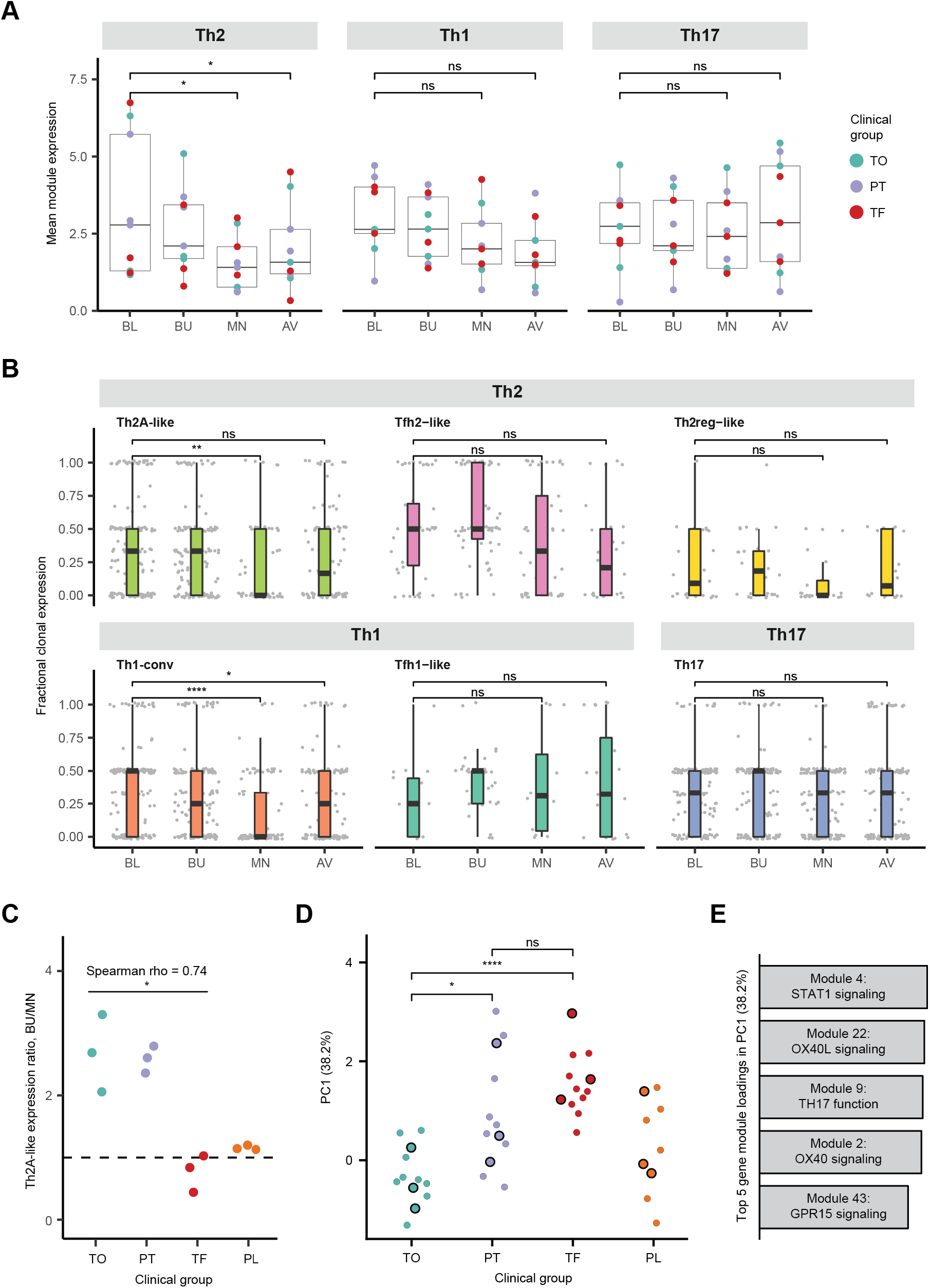
Th1 and Th2, but not Tfh, subsets are suppressed by OIT. **a,** Mean Th1, Th2 and Th17 module expression over time for treatment-group patients. Each data point represents the mean of all CD154+ cells for a given patient at a given time point. ‘*’ refers to an adjusted p-value of <0.05 by a Wilcoxon signed-rank test. **b**, Fractional expression of Th2, Th1, and Th17 modules within clonotypes of T helper subtypes over time. Fractional clonal expression is defined as the proportion of cells within each clonotype at a given time point expressing their respective module. Each data point represents the cells of an individual expanded clonotype from one patient at one timepoint. **c**, Degree of suppression in Th2A-like clones by clinical group. Ratio of mean Th2 module expression in Th2A-like clones from each patient was calculated between buildup (BU) and maintenance (MN). Spearman’s rho and p-value (‘*’ refers to p-value < 0.05) are from a Spearman correlation test between ratio and outcome within the treatment group (assigning TO as 2, PT as 1, and TF as 0). **d,** Principal component 1 (PC1) score of CD154+ cells by outcome. A principal components analysis was done using the 50 gene modules as features and all CD154+ cells at baseline as the input data. Each data point represents the mean PC1 score of all CD154+ cells from a single patient at a single timepoint. Black-outlined points represent the baseline timepoint. ‘*’ refers to an adjusted p-value of <0.05 by a Wilcoxon rank-sum test, “**” refers to adjusted p-value of <0.005, and “****” refers to adjusted p-value of <0.0005. **e,** Top 5 gene module loadings in PC1. Bar heights represent the magnitude of each contribution to PC1. Further details of each gene module are available in Supplementary Figures 3 and 4.

We then quantified the impact of OIT on gene module expression by individual clonotype in each of the T helper subtypes. This analysis allowed us to track the phenotypes of hundreds of individual clonal lineages over the course of treatment. To understand the stability of module expression over time within a clonotype, we calculated the “fractional clonal expression” for each clonotype by assigning it to the T helper subset in which it most frequently appeared, and then tabulating the proportion of cells that expressed the corresponding module (Th2, Th1, or Th17) at each time point (**Methods**). We found that Th1-conv and Th2A-like clonotypes had suppressed expression of Th1 and Th2 genes, respectively, at the maintenance time point compared to baseline. This suppression was consistent with an anergic state, characterized by decreased cytokine expression in response to stimulation, and was not detected in the placebo group (**Supplementary Figure** 9C). In contrast, we did not observe statistically significant changes in module expression at the clonotype-level in the Tfh1-like, Tfh2-like, Th2reg-like, or Th17 subsets (**Figure** 4B; **Supplementary Figure** 9C), suggesting that these populations are more refractory to modulation by OIT than Th1-conv and Th2A-like clonotypes. A lack of suppression of Th2A- like clonotypes at maintenance was associated with treatment failure (Spearman’s rho of 0.74 and unadjusted p-value of 0.02), although the degree of suppression did not differ between partial and full tolerance groups (**Figure** 4C). No statistically significant association was found in Th1-conv clonotypes (**Supplementary Figure** 9A).

### Non-Th2 inflammatory pathways at baseline are associated with clinical outcome

While a lack of Th2 suppression during OIT was associated with poor clinical outcome, its baseline expression was not predictive. To analyze immune signatures prior to the start of treatment, we performed PCA on gene module scores of all CD154+ cells at baseline. This approach allowed us to assess major axes of phenotypic variation among CD154+ cells at baseline and investigate whether any of these axes correlate with clinical outcome. We found a striking separation by outcome at all timepoints in the scores of the first principal component (PC1) alone, with high PC1 scores associated with poor clinical outcome (**Figure** 4D). The top gene modules enriched in PC1 were defined by markers of T cell activation and effector response such as OX40, OX40L, Th17 function, STAT1, and GPR15 (**Figure** 4E; **Supplementary Figure** 10). To investigate the cell types associated with this signature, we summarized PC1 scores and module expression in the six previously identified Th subtypes. Of these, Th1-conv and Th17 cells expressed the highest levels of PC1 (**Supplementary Figure** 11). Consistent with this observation, the frequencies of Th1-conv and Th17, but not Th2, cells were also lower in patients with favorable clinical outcome within the CD154+ compartment (**Supplementary Figure** 7). Interestingly, CD154+ cells not classified within any of the canonical CD4 T cell subtypes also showed outcome-dependent expression of modules associated with PC1 (**Supplementary Figure** 10; **Supplementary Figure** 12). These results indicate that a range of CD4 T cell phenotypes and inflammatory pathways may impact the likelihood of favorable responses to OIT.

## Discussion

In this study, we characterized peanut-reactive T helper cells from allergic patients undergoing OIT using single-cell RNA sequencing with paired TCR sequencing. These methods allowed us to identify patterns of expansion and TCR convergence among distinct peanut-reactive T helper subtypes and to longitudinally profile individual clonotypes throughout OIT. We found differential effects of OIT on T helper subtypes and a significant association at baseline between T cell phenotypes and clinical outcome, that is unmodulated by OIT. Our results add refinement to the transcriptomic-scale definitions of previously described subsets, reveal how clonotypes from these populations are affected during OIT, and provide additional insight into the substantial heterogeneity of peanut allergic patients.

Among sorted CD154+ and CD137+ T helper cells, we identified six subtypes of highly clonal peanut-reactive T helper cells with Th1, Th2, and Th17 signatures. Of these subtypes, the Th2A- like, Tfh2-like, and Th2reg-like cells correspond well to the previously described Th2A, Tfh13, and deviated Treg populations in food allergy^8–10,35^. We show here for the first time that these subsets have distinct TCR repertoires and that some are enriched in highly similar TCR sequences. Our results add resolution to a previous study showing that distinct repertoires exist between CD154+ and CD137+ cells^27^. This segregation of TCR repertoires strongly suggests that the subsets represent distinct lineages rather than transient phenotypes, and the phenomenon of TCR convergence hints at the skewing of T cell state due to epitope interactions or epitope-associated factors^41–43^. We did not detect significant TCR convergence in the Th2A-like and the Th2reg-like subsets, which could be due to the higher diversities of repertoires in these subsets that would require deeper sampling to detect any convergence.

We also showed, for the first time, how these three Th2 subsets, as well as Th1 and Th17 cells, responded over the course of OIT. Globally, we found that OIT induced a transient reduction of the frequency of CD154+ T cells and the mean expression of Th2 signatures in response to peanut antigen stimulation. However, we also found that the TCR repertoires of peanut-reactive Th2 cells were stable over time, suggesting that OIT acts predominantly via suppression of functional phenotypes rather than by clonal deletion or TCR-biased sequestration away from the periphery. This result corroborates two previous studies that reported the emergence of anergic signatures in peanut-specific T cells over time in OIT, and provides insight into previous reports of decreases in circulating Th2 frequency following OIT^8,18,19,22,23^. Furthermore, we observed OIT-induced suppression among Th2A-like and Th1-conv, but not Tfh-like, clonotypes. Finally, we observed that the suppression of Th2 module expression in Th2A-like clonotypes was associated with clinical outcome. Our findings indicate that OIT is successful in modulating only a subset of peanut-reactive T cells, and that the T cells most responsible for Ig class-switching and B cell help may be the least altered by treatment, highlighting the difficulty of achieving a sustained change in clinical outcome.

With respect to therapeutic outcomes, we found that an unsupervised composite score of all gene modules, derived using only data from cells isolated before treatment (baseline), corresponded strongly with treatment failure and was not modulated by treatment. This score was driven largely by markers of T cell activation such as *OX40, OX40L*, and *STAT1*, as well as Th1 and Th17 genes (**Supplementary Figure** 10C). We surmise that high levels of baseline T cell activation could limit the effectiveness of OIT due to increased inflammation or altered gastrointestinal permeability (potentially triggered by Th17 responses^44^). Assessments of genomic or immunologic features associated with clinical outcomes in OIT are scarce, but Th17 cells have been reported to play a role in atopic disease, with some preliminary evidence suggesting that they are modulated by OIT^26,45–47^. Similarly, OX40 and OX40L have also been implicated in atopic dermatitis and asthma, and represent a possible therapeutic target^48,49^.

Moreover, while some of the top gene modules in the composite score were highly enriched among Th1 and Th17 subsets (e.g., the OX40L module), many were also expressed in other compartments of CD154+ or CD137+ cells (**Supplementary Figure** 10), including CD154+ cells not classified as any of the Th subtypes. Such was the case for the GPR15 and STAT1 signaling modules (**Supplementary Figure** 12). GPR15 has been highlighted as a colon-homing receptor in CD4+ T cells, and STAT1 (along with GBP4 and GBP1, also included in the same module) is associated with response to interferon^50,51^. Taken together, these results could suggest that altered gastrointestinal permeability and inflammatory responses in diverse combinations of peanut-reactive T cells may influence the likelihood of responding favorably to OIT.

Tregs have been described in some studies as a correlate of favorable clinical outcome in OIT^20,21^. While we detected a strong and sustained expression of Treg markers among peanut-reactive CD137+ cells, we actually saw a moderate decrease in the frequency of CD137+ cells over the course of OIT (**Supplementary Figure** 1B). In addition, although *IL10* in both CD137+ and CD154+ was transiently induced, there was neither a sustained increase in Treg expression among CD137+ cells over OIT, as measured by the average level of expression of the Treg module or *FOXP3* and *IL10* (**Extended Data Figure** 3), nor a correlation of *IL10* or *FOXP3* with clinical outcome. Finally, we did not see evidence for the induction of new peanut-reactive Treg clonotypes during OIT, as TCR repertoires of CD137+ cells remained stable over time (**Extended Data Figure** 2). Discrepancies between our results and those of prior studies could reflect differences in stimulation conditions and strategies for identifying antigen-specific Tregs, and they motivate further efforts towards elucidating the role of peanut-reactive Tregs in OIT.

The methods we used in this study combine FACS-based enrichment of activated T cells with single-cell RNA-Seq and TCR sequencing as a framework for profiling antigen-reactive T cells without the use of tetramer reagents. By enriching peanut-reactive T cells based on CD154 and CD137 expression, it is likely that our data include some fraction of non-specific activated T cells. By integrating data on TCR sequences, however, we identified T cell states that were associated with both clonal expansion and peanut-antigen activation, thereby minimizing the effects of non-specifically activated T cells. We believe this framework could be used to identify likely antigen-reactive T cells in other disease contexts.

We believe this work has implications for the study of human T cell biology as well as mechanisms of OIT. First, the methodology implemented here provides a framework for the design and analysis of paired TCR and transcriptome data of antigen-reactive T cells, and this substantial dataset of human single-cell data provides a useful reference for future studies. Using this framework, we detected significant heterogeneity within the peanut-reactive CD154+ T cell compartment and highlighted potential roles for TCR-epitope interactions in skewing T cell phenotype. Second, our data have revealed several features of OIT that merit further investigation. Based on our data, OIT does not appear to delete peanut-reactive Th2 clones; these findings point to selective clonal suppression, rather than deletion, as a major mechanism of OIT and highlight why sustained tolerance may be difficult to achieve. Furthermore, we found that failure to respond to OIT was reflected in a broad baseline activation signature, highly expressed in Th17 and other T cells, that was resistant to modulation by OIT. In the future, prospective OIT studies could evaluate this signature as a predictor of treatment success. In summary, we used single-cell RNA-Seq and TCR clonotyping to reveal a complex set of highly distinct T helper cell phenotypes, beyond effector Th2, that are relevant to the efficacy of OIT. Future therapeutic modalities that either target these diverse phenotypes and inflammatory pathways, such as Tfh, Th17, OX40-OX40L, or that appreciably delete peanut-specific Th2A cells, may be more likely to promote sustained tolerance in food allergy than allergen-based approaches alone.

## Data Availability

FASTQ file format data related to human samples will be available through dbGaP under accession number phs001897.v1.p1. Processed gene expression and associated TCR clonotype data will be available through GEO under accession number GSE158667. Processed data files and associated meta data tables for Figures 1-4 will be made available on https://github.com/mitlovelab/, or upon request.

## Author Contributions

W.G.S. and J.C.L. conceptualized the study. W.G.S. conducted the clinical trial (NCT01750879). B.M., A.A.T., B.R., and P.M.P. conducted experiments. B.M., A.A.T., D.M.M., and N.P.S. analyzed the data. T.M.G. and J.H.G. conducted experiments to develop and validate methods. B.M., A.A.T., B.R., D.M.M., W.G.S., and J.C.L. wrote and edited the manuscript.

## Acknowledgements

We would like to thank our patients and their families who generously gave their time and participation, as well as Lauren Tracy, Colby Rondeau, Christine Elliot and Leah Hayden, the clinical coordinators of this study. We would also like to thank Sarita U. Patil and Yamini V. Virkud for fruitful discussions. The clinical work was performed in the Harvard Clinical and Translational Science Center supported by grants 1UL1TR001102 and 8UL1TR000170 from the National Center for Advancing Translational Sciences, and 1UL1RR025758 from the National Center for Research Resources. In addition, we thank our colleagues at the MGH Department of Pathology Flow and Image Cytometry Research Core for their help with cell sorting. The Flow Core obtained funding from the National Institutes of Health Shared Instrumentation program (1S10OD012027-01A1, 1S10OD016372-01, 1S10RR020936-01, and 1S10RR023440-01A1). This work was supported in part by the Koch Institute Support (core) NIH Grant P30-CA14051 from the National Cancer Institute, as well as the Koch Institute - Dana-Farber/Harvard Cancer Center Bridge Project. This work was also supported by the Food Allergy Science Initiative at the Broad Institute and the NIH (5P01AI039671, 5U19AI089992, U19AI095261).

## Methods

### Patients

Peanut-allergic individuals age 7 years and older were enrolled in a peanut OIT trial (NCT01750879) at the Food Allergy Center at Massachusetts General Hospital. All subjects were recruited with informed consent, and the study was approved by the Institutional Review Board of Partners Healthcare (protocol 2012P002153). Study participants with a previous diagnosis of peanut allergy, a history of peanut-induced reactions consistent with immediate hypersensitivity, and confirmatory peanut- and Ara h 2–specific serum IgE concentrations (peanut-specific IgE > 5 kU/L, Ara h 2–specific IgE > 0.35 kU/L; ImmunoCAP; Thermo Fisher, Waltham, MA) underwent a double-blind placebo-controlled food challenge (DBPCFC).

Increasing peanut protein doses were administered every 20 minutes to a maximum dose of 300 mg according to the following schedule: 3, 10, 30, 100, and 300 mg, for a cumulative total of 443 mg. Patients who had an objective allergic reaction during the challenge were eligible for inclusion in the study.

### Oral immunotherapy (OIT) study

The main objective of this phase I/II, double-blind placebo-controlled, interventional study was to provide safety and mechanistic data on OIT for people with IgE-mediated peanut allergy. Enrolled patients were randomized to receive either treatment (peanut flour) or placebo (roasted oat flour) at a ratio of 3:1. Treatment consisted of a modified-rush protocol, followed by a build-up phase lasting for 44 weeks or when the patient reached 4000mg, whichever came first. Treatment dose was administered daily, and dosing escalation was incremental, based on previous OIT studies^8^, occurring every two weeks. After the buildup phase, patients entered a maintenance phase in which treatment was continued at the top tolerated dose for each patient for 12 weeks. Finally, patients underwent an avoidance phase, 12 weeks off therapy while strictly avoiding dietary peanut protein, in order to assess the durability of any desensitization resulting from OIT. During each phase of the study, a blood sample was taken, for four samples total per patient: 2 weeks prior to the start of treatment at baseline, 14 weeks into the buildup phase, 8 weeks into the maintenance phase, and 8 weeks into the avoidance phase.

Clinical assessments were made by double-blind placebo-controlled food challenge at baseline (DBPCFC1), at the end of 12 weeks of maintenance therapy (DBPCFC2), and at the end of 12 weeks of avoidance^21^ (DBPCFC3). Clinical outcomes were defined as: treatment failure (failure to achieve the minimum maintenance dose (600 mg) of peanut protein by 12 months, or an eliciting dose less than 1443 mg at DBPCFC2, or less than 443mg at DBPCFC3, OR less than 10-fold more than at DBPCFC1); partial tolerance (eliciting dose less than 4430mg at DBPCFC3 but at least 443 mg AND more than 10-fold more than at DBPCFC1); and tolerance (ingestion of 4430 mg of peanut protein at DBPCFC3 without symptoms).

### Cell purification and sorting

After a blood sample was collected, PBMCs were isolated by density gradient centrifugation (Ficoll-Paque Plus; GE Healthcare) and cryopreserved in FBS with 10% DMSO. After the study was completed, for each of the 12 patients, PBMCs from all four time points (15-30 x 10^6^ PBMCs per timepoint) were simultaneously thawed, washed with PBS, and cultured in AIM-V medium (Gibco) with 100 µg/ml peanut protein extract for 20h, at a density of 5 x 10^6^ PBMCs in 1 mL medium per well in 24-well plates. Peanut protein extract was prepared by agitation of defatted peanut flour (Golden Peanut and Tree Nuts, Alpharetta, GA) with PBS, centrifugation, and sterile-filtering. Endotoxin concentration in the peanut protein extract was assessed to be 6 EU/mg, using a LAL Endotoxin Quantitation kit (Thermo Fisher; cat. no. 88282). This is lower than the concentration in commercially available endotoxin-depleted preparations of the purified peanut proteins Ara h 1 and Ara h 2 (Indoor Biotechnologies; LTN-AH1-1 and LTN-AH2-1). Furthermore, the endotoxin concentration in the PBMC cultures with peanut protein extract was 0.6 EU/ml, which is comparable to the endotoxin limit for eluates from medical devices (0.5 EU/ml) as determined by the FDA^52^. Anti-CD154-PE antibody (BD Biosciences; clone TRAP1) was added to the cultures at a 1:50 dilution (20 µl/well) for the last 3h. After harvesting, the cells were labeled with anti-CD3-AF700 (BD Biosciences; UCHT1), anti-CD4-APC-Cy7 (BD Biosciences; RPA-T4), anti-CD45RA-PE-Cy7 (BD Biosciences; HI100), anti-CD154-PE (BD Biosciences; TRAP1), anti-CD137-APC (BD Biosciences; clone 4B4-1), and Live/Dead Fixable Blue stain (Thermo Fisher; cat. no. L23105). Cells were then sorted with a FACSAria Fusion instrument (BD Biosciences). Cells were gated as live singlet CD3+CD4+CD45RA- and then sorted as either CD154+CD137+/− (referred to as “CD154+”), CD154-CD137+ (“CD137+”), or CD154-CD137-.

### Single-cell RNA-Seq

Sorted subsets of CD4 memory T cells were processed for single-cell RNA sequencing using the Seq-Well platform as previously described^31^. A portion of each cDNA library was reserved for paired TCRα/β enrichment. The rest was barcoded and amplified using the Nextera XT kit and sequenced on the Illumina NovaSeq.

Raw read processing was performed as in Macosko et al^53^. Briefly, sequencing reads were aligned to the ‘hg38’ reference human genome, collapsed by unique molecular identifier (UMI), and counted to obtain a digital gene expression matrix of cells versus genes. These counts were then filtered to exclude any cells with fewer than 1000 genes or 2000 UMIs and normalized by library size per cell and a log2 transformation. For the rare T helper subsets analysis which required more cells, a filter of 500 genes and 1000 UMIs was used.

### Paired single-cell TCRαβ sequencing

Paired TCR sequencing was performed according to Tu et al^32^. Briefly, following cDNA amplification, biotinylated capture probes for human TRAC and TRBC regions were annealed to cDNA. Magnetic streptavidin beads were used to enrich the bound TCR sequences, which were then further amplified using human V-region primers and prepared for sequencing using Nextera sequencing handles. Libraries were sequenced on an Illumina MiSeq using 150bp-length reads.

TCR sequencing reads were preprocessed according to Tu et al^32^. In short, reads were mapped to TCRV and TCRJ IMGT reference sequences via IgBlast, and V and J calls with “strong plurality” (wherein the ratios of the most frequent V and J calls to the second most frequent calls were at least 0.6) were retained. CDR3 sequences were called by identifying the 104-cysteine and 118-phenylalanine according to IMGT references and translating the amino acid sequences in between those residues. Processed TCR sequences were then paired with the single-cell transcriptome data via the cell barcodes.

### Visualization of single-cell RNA-Seq data

Visualization of single-cell transcriptomes was done with UMAP^54^ (uniform manifold approximation and projection) with the Python package “scanpy”. Prior to visualization, the normalized gene expression data was transformed using a standard “regress-out” approach to mitigate batch effects: a multiple linear regression was performed on all genes with two covariates that could be batch-associated (numbers of transcripts per cell, and percent of transcripts aligning to the mitochondrial chromosome). The residuals from this regression were taken as the transformed data. Next, a principal components analysis was performed, and the top 10 components were used to generate a UMAP visualization.

### Gene module discovery

Coexpressed gene modules were generated based on a sparse PCA approach described by Witten et al and implemented in the R package “PMA”^33^. This unsupervised method employs an L1 norm penalty on loadings in each component to introduce sparsity. Prior to running sparse PCA, the gene expression matrix was randomly downsampled to have an equal number of cells from all samples, to prevent the results from being skewed by a subset of the samples. Genes were filtered down to the union of immune genes (defined by the set of gene lists available on ImmPort at https://www.immport.org/shared/genelists) and the variable genes in the dataset (defined using the R package “Seurat”). Finally, the gene expression data was scaled with respect to genes, and sparse PCA was run using the command “SPC”. Gene module scores were calculated as the scaled gene expression input matrix multiplied by the outputted loadings matrix “v”. The first 50 gene modules were retained for downstream analysis.

Cells were classified as “expressing” or “not expressing” a module using a simple thresholding strategy. The distribution of module scores of CD154-CD137-cells was used as a negative control, and a threshold was set at the point where 0.2% of CD154-CD137-cells were in the positive population. Cells with a module score above the threshold were labeled as “expressing” that module. Modules in which at least 60% of the expressing cells were from a single patient were removed from downstream analysis.

### Identification of T helper subtypes

All CD154+ and CD137+ cell transcriptomes were classified as Th1, Th2, or Th17 using the criteria for module expression detailed above (see **Gene module discovery**) for the Th1, Th2, and Th17 gene modules. If a cell expressed more than one Th module, it was assigned to the module with the highest z-score (compared to the distribution of all CD154+ and CD137+ cells). Then, each individual Th class (Th1, Th2, and Th17 cells) was separately visualized by UMAP and clustered by Louvain clustering using the R package ‘Seurat’.

### Distance analysis of TCR sequences

Pairwise distance of TCRβCDR3 sequences was evaluated using the TCRdist method published by Dash et al^38^. Briefly, for two TCRβ CDR3 amino acid sequences of the same length, each residue position was compared, and a penalty was assessed for every mismatch. The penalty for two different amino acid residues i and j was assessed using the BLOSUM62 matrix and was defined as min(4 – BLOSUM62[i, j], 4). Each substitution thus incurred a penalty between 1 and 4. The overall distance between two CDR3s was calculated as the sum of penalties at all positions. In the case of two CDR3s of unequal length, the sequences were aligned in all possible ways and the minimum overall penalty was taken, with each gap incurring a penalty of 8.

### Likelihood-based association between TCR and T helper subtype

Likelihood-based analysis was used to determine the tightness of association between T helper subset and TCRβ CDR3 sequence. A log-likelihood ratio was defined as log_2_(P/P_0_), where P was the probability of two cells being of the same T helper subset if they were drawn randomly from all cells sharing the same TCRβ CDR3 sequence (without replacement), and P_0_ was the probability of two cells being of the same T helper subset if they were drawn randomly from all cells. P_0_ represents the prior probability without the constraint of TCR information; thus, the ratio P/P_0_ represents the gain in likelihood due to the knowledge of TCR sequence. This analysis was constrained to consider all pairs of cells within the same patient.

### Longitudinal analysis of clonotypes

Temporal analysis of individual clonotypes over the course of OIT involved two analyses: 1) obtaining the distribution of timepoints at which each clonotype was detected, and 2) assessing module expression of clonotypes within T helper subtypes. For the former, CD154+ or CD137+ clones were filtered to those with at least 4 cells in that sorted subset. Then, the timepoints covered by the cells was tabulated and the clonotype was classified as having a specific temporal pattern (e.g. “BL, BU, AV”). For the latter, clonotypes were filtered down to those with at least 2 cells in the combined CD154+ and CD137+ compartments. Each clonotype was then assigned to a T helper subtype (or no subtype) based on the most frequent T subtype that its cells mapped to (see **Identification of T helper subtypes**). At each timepoint, the fraction of cells within each clonotype expressing the relevant module (Th1, Th2, or Th17) was counted, relative to the total number of cells of that clonotype at that timepoint. Fractional expression was used instead of module scores in order to normalize for clonotype- or patient-driven differences in dynamic range of module expression.

### Baseline signature of all modules using PCA

Principal component analysis was used to identify broad immune signatures associated with clinical outcome. Mean module scores of the 50 gene modules (minus the 7 modules associated with a single patient; see **Gene module discovery**) were computed for each patient at each timepoint. Averages at baseline were used to compute the principal components. The first principal component (“PC1”), or the component explaining the largest amount of variance, was then applied to module averages of other timepoints.

### Spearman correlation of treatment outcome and module expression

To investigate whether treatment outcome was correlated with expression of modules enriched in PC1, we assigned numerical values to each of the outcomes (TO as 2, PT as 1, and TF as 0) to represent an ordinal relationship between the outcomes. Spearman correlation between mean module expression (by patient and cell subset) and outcomes was calculated. Corresponding unadjusted p-values were reported.

### Statistical analyses

All statistical tests were performed as two-sided tests unless otherwise noted. Box plots were plotted with the standard visualization of 25^th^ and 75^th^ percentile for the lower and upper hinges, and at most 1.5 times the interquartile range for the whisker lengths.

### Code availability

R, python, and Matlab scripts for processing TCR sequencing data and generating all analyses, as well as all updates, will be made available on https://github.com/mitlovelab or upon request.

---

**Extended Data 1.**
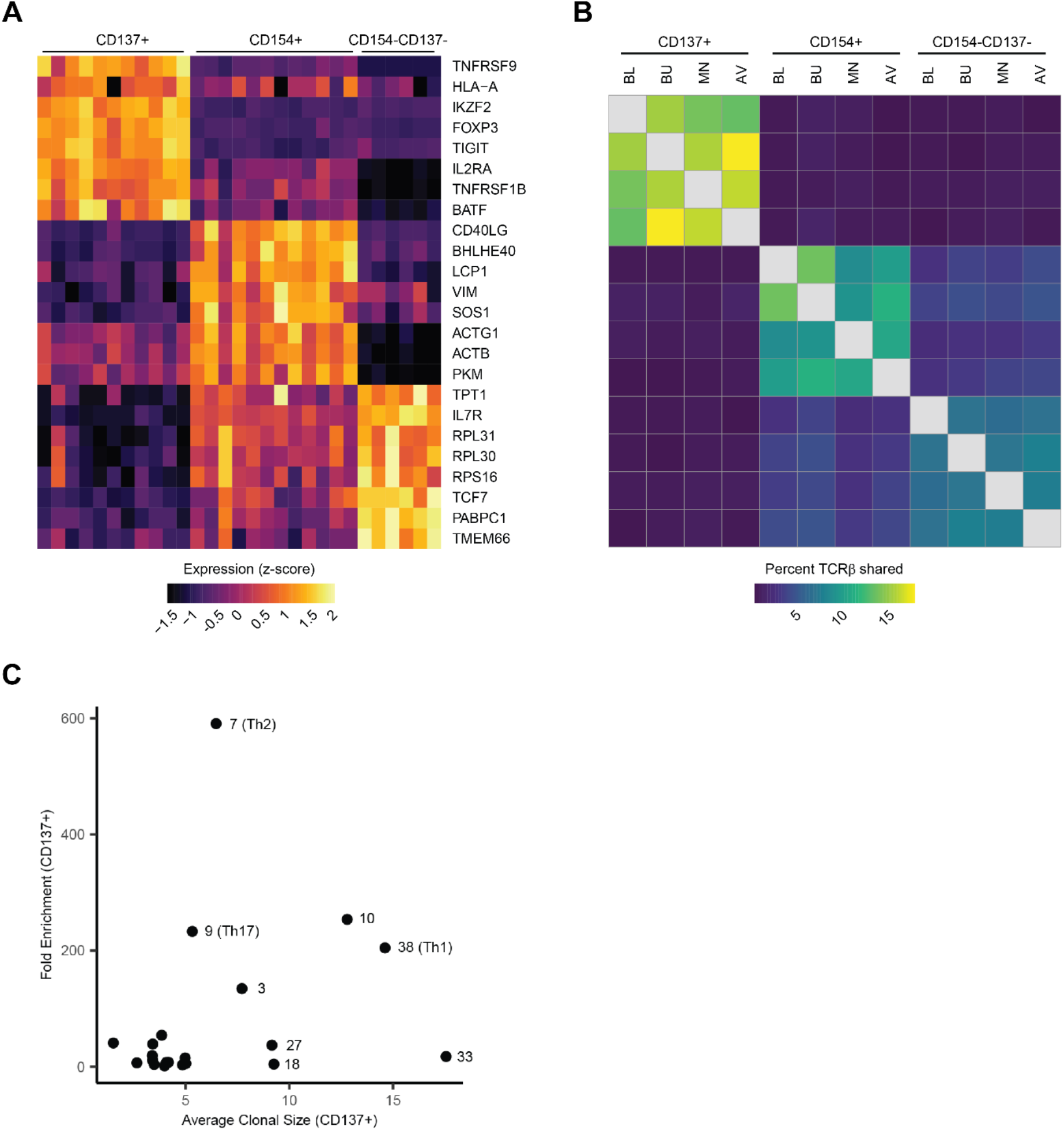
**a,** Top differentially expressed genes between the sorted subsets. Each column represents the scaled average gene expression of cells from a single patient. Genes were selected using ROC test. **b,** Percent of TCRβ sequences shared between time points and all sorted subsets. ‘Percent shared’ is defined as the number of unique TCRβ sequences detected in both conditions, divided by the geometric mean of the number of unique TCRβ sequences in each of the two conditions. Sequences from all treatment-group patients were pooled. Patients without recovered TCR sequences from all three sorted subsets were excluded from the analysis. **c,** Mean clonal size and fold-change in mean module scores (compared to module-expressing CD154-CD137-cells) in CD137+ cells expressing each gene module. Cells were classified as ‘expressing’ each module or not, relative to background expression (**Methods**). Clonal size was calculated with respect to all cells in the dataset.

**Extended Data 2.**
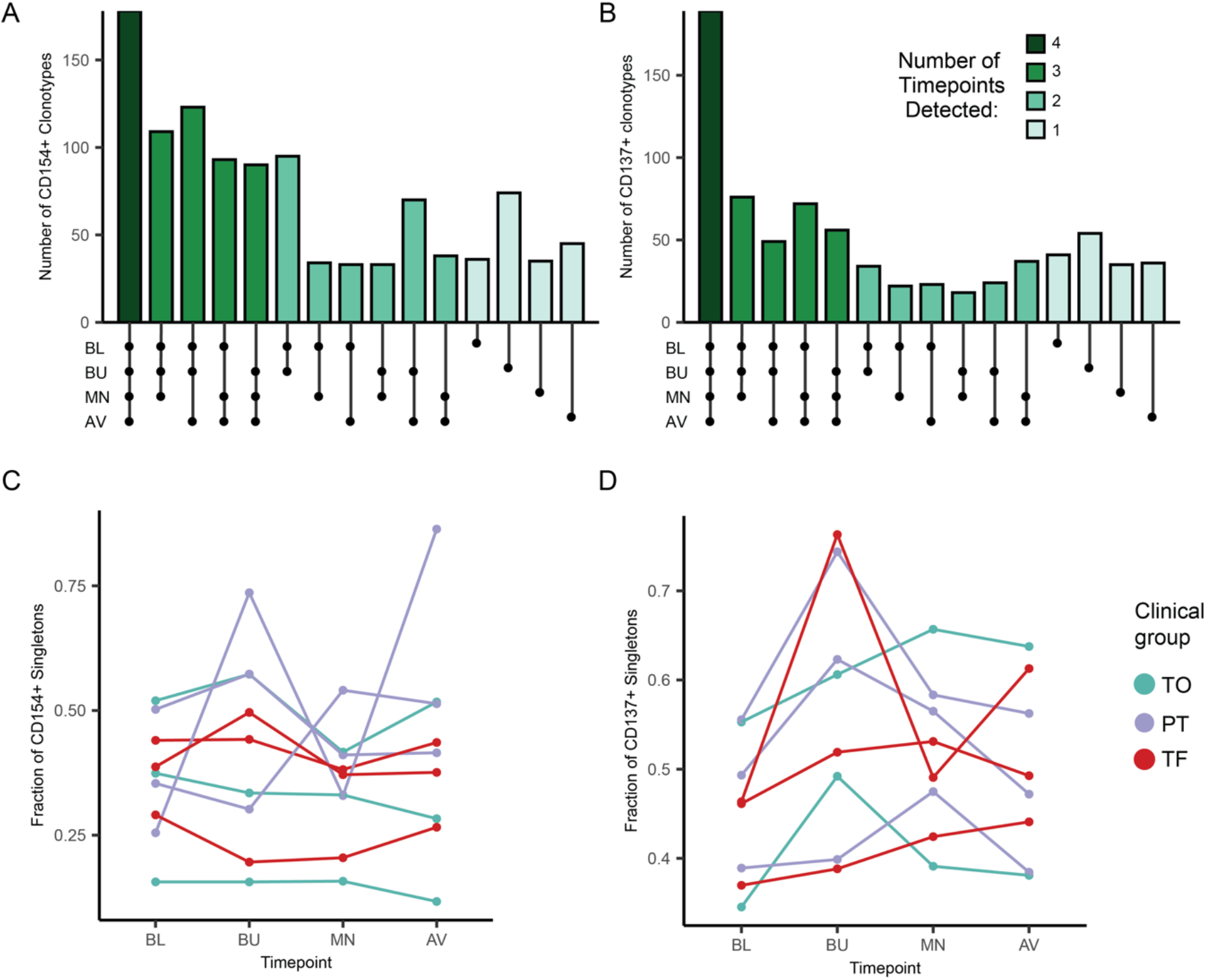
**a-b**, Number of TCRβ clonotypes detected at every possible combination of timepoints in (**a**) CD154+ and (**b**) CD137+ cells. Clones from all treatment-group patients with at least 4 cells were included. **c**-**d**, Fraction of singletons (clonotypes with clonal size of one) detected within each patient and at each timepoint in (**c**) CD154+ and (**d**) CD137+ cells.

**Extended Data 3.**
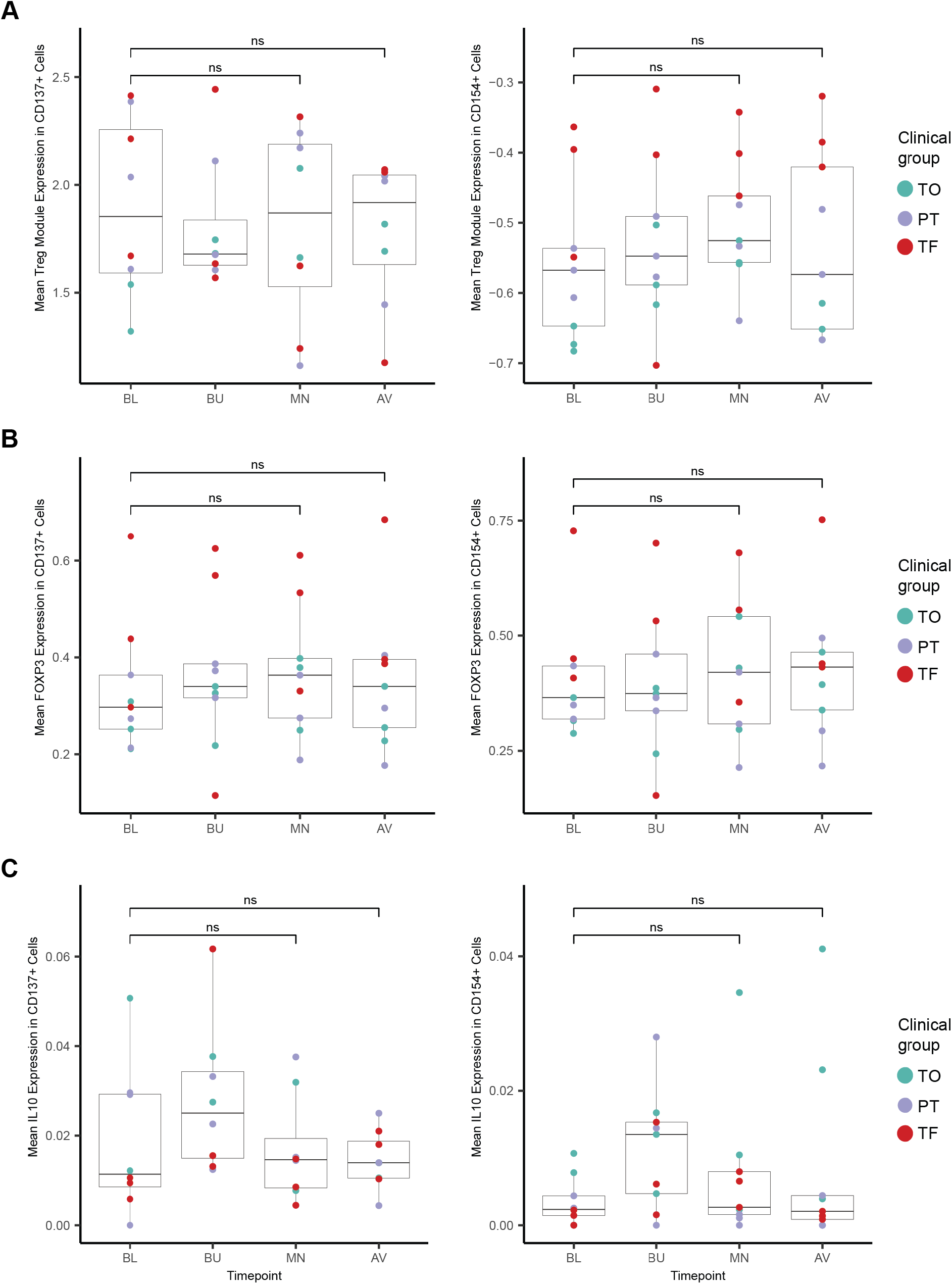
**a**, Average expression of Treg module (module 1) by patient and timepoint within CD137+ (left) and CD154+ (right) cells. **b**, Average expression of *FOXP3* by patient and timepoint within CD137+ (left) and CD154+ (right) cells. **c**, Average expression of *IL10* by patient and timepoint within CD137+ (left) and CD154+ (right) cells. Adjusted p-values calculated by Wilcoxon rank-sum test (**a**-**c**).

## References

1. Gupta, R. S. et al. The prevalence, severity, and distribution of childhood food allergy in the United States. Pediatrics 128, (2011).

2. Sicherer, S. H. & Sampson, H. A. Food allergy: A review and update on epidemiology, pathogenesis, diagnosis, prevention, and management. J. Allergy Clin. Immunol. 141, 41–58 (2018).

3. Vickery, B. P. et al. AR101 Oral Immunotherapy for Peanut Allergy. N. Engl. J. Med. 379, 1991–2001 (2018).

4. Patil, S. U. et al. Peanut oral immunotherapy transiently expands circulating Ara h 2–specific B cells with a homologous repertoire in unrelated subjects. J. Allergy Clin. Immunol. 136, 125–134.e12 (2015).

5. Vickery, B. P., Chin, S. & Burks, A. W. Pathophysiology of Food Allergy. Pediatr. Clin. North Am. 58, 363–376 (2011).

6. Prussin, C., Yin, Y. & Upadhyaya, B. TH2 heterogeneity: Does function follow form? J. Allergy Clin. Immunol. 126, 1094–1098 (2010).

7. Sampath, V. & Nadeau, K. C. Newly identified T cell subsets in mechanistic studies of food immunotherapy. Journal of Clinical Investigation 129, 1431–1440 (2019).

8. Wambre, E. et al. A phenotypically and functionally distinct human TH2 cell subpopulation is associated with allergic disorders. Sci. Transl. Med. 9, eaam9171 (2017).

9. Gowthaman, U. et al. Identification of a T follicular helper cell subset that drives anaphylactic IgE. Science (80-. ). 365, eaaw6433 (2019).

10. Mitson-Salazar, A. et al. Hematopoietic prostaglandin D synthase defines a proeosinophilic pathogenic effector human T H 2 cell subpopulation with enhanced function. J. Allergy Clin. Immunol. 137, 907–918.e9 (2016).

11. Ruiter, B. et al. Expansion of the CD4+ effector T-cell repertoire characterizes peanut-allergic patients with heightened clinical sensitivity. J. Allergy Clin. Immunol. 145, 270–282 (2020).

12. Haribhai, D. et al. A requisite role for Induced Regulatory T cells in Tolerance Based on Expanding Antigen Receptor Diversity. Immunity 35, 109–122 (2011).

13. Chinthrajah, R. S. et al. Sustained outcomes in oral immunotherapy for peanut allergy (POISED study): a large, randomised, double-blind, placebo-controlled, phase 2 study. Lancet 394, 1437–1449 (2019).

14. Blumchen, K. et al. Oral peanut immunotherapy in children with peanut anaphylaxis. J. Allergy Clin. Immunol. 126, 83–91.e1 (2010).

15. Vickery, B. P. et al. Sustained unresponsiveness to peanut in subjects who have completed peanut oral immunotherapy. J. Allergy Clin. Immunol. 133, 468–475.e6 (2014).

16. Blumchen, K. et al. Efficacy, Safety, and Quality of Life in a Multicenter, Randomized, Placebo-Controlled Trial of Low-Dose Peanut Oral Immunotherapy in Children with Peanut Allergy. J. Allergy Clin. Immunol. Pract. 7, 479–491.e10 (2019).

17. Kim, E. H. et al. Sublingual immunotherapy for peanut allergy: clinical and immunologic evidence of desensitization. J. Allergy Clin. Immunol. 127, 640–6.e1 (2011).

18. Ryan, J. F. et al. Successful immunotherapy induces previously unidentified allergen-specific CD4+ T-cell subsets. Proc. Natl. Acad. Sci. U. S. A. 113, E1286–E1295 (2016).

19. Frischmeyer-Guerrerio, P. A. et al. Mechanistic correlates of clinical responses to omalizumab in the setting of oral immunotherapy for milk allergy. J. Allergy Clin. Immunol. 140, 1043–1053.e8 (2017).

20. Syed, A. et al. Peanut oral immunotherapy results in increased antigen-induced regulatory T-cell function and hypomethylation of forkhead box protein 3 (FOXP3). J. Allergy Clin. Immunol. 133, 500–510.e11 (2014).

21. Varshney, P. et al. A randomized controlled study of peanut oral immunotherapy: Clinical desensitization and modulation of the allergic response. J. Allergy Clin. Immunol. 127, 654–660 (2011).

22. Weissler, K. A. et al. Identification and analysis of peanut-specific effector T and regulatory T cells in children allergic and tolerant to peanut. J. Allergy Clin. Immunol. 141, 1699–1710.e7 (2018).

23. Wang, W. et al. Transcriptional changes in peanut-specific CD4+ T cells over the course of oral immunotherapy. Clin. Immunol. 219, 108568 (2020).

24. Wambre, E. Effect of allergen-specific immunotherapy on CD4+ T cells. Current Opinion in Allergy and Clinical Immunology 15, 581–587 (2015).

25. Tordesillas, L. & Berin, M. C. Mechanisms of Oral Tolerance. Clinical Reviews in Allergy and Immunology 55, 107–117 (2018).

26. Chiang, D. et al. Single-cell profiling of peanut-responsive T cells in patients with peanut allergy reveals heterogeneous effector T H 2 subsets. J. Allergy Clin. Immunol. 141, 2107–2120 (2018).

27. Bacher, P. et al. Regulatory T Cell Specificity Directs Tolerance versus Allergy against Aeroantigens in Humans. Cell 167, 1067–1078.e16 (2016).

28. Aguet, F. et al. Genetic effects on gene expression across human tissues. Nature 550, 204–213 (2017).

29. Kunnath-Velayudhan, S. et al. Transcriptome Analysis of Mycobacteria-Specific CD4 + T Cells Identified by Activation-Induced Expression of CD154. J. Immunol. 199, 2596–2606 (2017).

30. Commandeur, S. et al. Clonal Analysis of the T-Cell Response to In Vivo Expressed Mycobacterium tuberculosis Protein Rv2034, Using a CD154 Expression Based T-Cell Cloning Method. PLoS One 9, e99203 (2014).

31. Gierahn, T. M. et al. Seq-Well: portable, low-cost RNA sequencing of single cells at high throughput. Nat. Methods 14, 395–398 (2017).

32. Tu, A. A. et al. TCR sequencing paired with massively parallel 3′ RNA-seq reveals clonotypic T cell signatures. Nat. Immunol. 20, 1692–1699 (2019).

33. Witten, D. M., Tibshirani, R. & Hastie, T. A penalized matrix decomposition, with applications to sparse principal components and canonical correlation analysis. Biostatistics 10, 515–534 (2009).

34. Kim, C. J. et al. The Transcription Factor Ets1 Suppresses T Follicular Helper Type 2 Cell Differentiation to Halt the Onset of Systemic Lupus Erythematosus. Immunity 49, 1034–1048.e8 (2018).

35. NovalRivas, M. et al. Regulatory T cell reprogramming toward a Th2-Cell-like lineage impairs oral tolerance and promotes food allergy. Immunity 42, 512–523 (2015).

36. Velu, V. et al. Induction of Th1-Biased T Follicular Helper (Tfh) Cells in Lymphoid Tissues during Chronic Simian Immunodeficiency Virus Infection Defines Functionally Distinct Germinal Center Tfh Cells. J. Immunol. 197, 1832–1842 (2016).

37. James, K. R. et al. Distinct microbial and immune niches of the human colon. Nat. Immunol. 21, 343–353 (2020).

38. Dash, P. et al. Quantifiable predictive features define epitope-specific T cell receptor repertoires. Nature 547, 89–93 (2017).

39. Hayes, S. M., Li, L. Q. & Love, P. E. TCR signal strength influences αβ/γδ lineage fate. Immunity 22, 583–593 (2005).

40. Daniels, M. A. & Teixeiro, E. TCR Signaling in T Cell Memory. Front. Immunol. 6, 617 (2015).

41. Snook, J. P., Kim, C. & Williams, M. A. TCR signal strength controls the differentiation of CD4+ effector and memory T cells. Sci. Immunol. 3, (2018).

42. Tubo, N. J. et al. Single naive CD4+ T cells from a diverse repertoire produce different effector cell types during infection. Cell 153, 785–796 (2013).

43. Kotov, D. I. et al. TCR Affinity Biases Th Cell Differentiation by Regulating CD25, Eef1e1, and Gbp2. J. Immunol. 202, 2535–2545 (2019).

44. Stockinger, B. & Omenetti, S. The dichotomous nature of T helper 17 cells. Nature Reviews Immunology 17, 535–544 (2017).

45. Choy, D. F. et al. TH2 and TH17 inflammatory pathways are reciprocally regulated in asthma. Sci. Transl. Med. 7, 301ra129 (2015).

46. Hirahara, K. & Nakayama, T. CD4+ T-cell subsets in inflammatory diseases: Beyond the Th1/Th2 paradigm. International Immunology 28, 163–171 (2016).

47. Luce, S., Chinthrajah, S., Lyu, S.-C., Nadeau, K. C. & Mascarell, L. Th2A and Th17 cell frequencies and regulatory markers as follow-up biomarker candidates for successful multifood oral immunotherapy. Allergy 75, 1513–1516 (2020).

48. Guttman-Yassky, E. et al. GBR 830, an anti-OX40, improves skin gene signatures and clinical scores in patients with atopic dermatitis. J. Allergy Clin. Immunol. 144, 482–493.e7 (2019).

49. Seshasayee, D. et al. In vivo blockade of OX40 ligand inhibits thymic stromal lymphopoietin driven atopic inflammation. J. Clin. Invest. 117, 3868–3878 (2007).

50. Adamczyk, A. et al. Differential expression of GPR15 on T cells during ulcerative colitis. JCI insight 2, (2017).

51. Ovadia, A., Sharfe, N., Hawkins, C., Laughlin, S. & Roifman, C. M. Two different STAT1 gain-of-function mutations lead to diverse IFN-γ-mediated gene expression. npj Genomic Med. 3, 23 (2018).

52. Guidance for Industry: Pyrogen and Endotoxins Testing: Questions and Answers | FDA. Available at: https://www.fda.gov/regulatory-information/search-fda-guidance-documents/guidance-industry-pyrogen-and-endotoxins-testing-questions-and-answers. (Accessed: 2nd March 2021)

53. Macosko, E. Z. et al. Highly Parallel Genome-wide Expression Profiling of Individual Cells Using Nanoliter Droplets. Cell 161, 1202–1214 (2015).

54. McInnes, L., Healy, J. & Melville, J. UMAP: Uniform Manifold Approximation and Projection for Dimension Reduction. (2018).

