## Supplement for "Peanut oral immunotherapy differentially suppresses clonally distinct subsets of T helper cells"

Supplementary Table 1. Patient demographics and baseline characteristics.

| Patient ID | Treatment Group | Age Range | Gender | Race | Peanut IgE (kU/L) | Peanut IgG4 (kU/L) | Total IgE (kU/L) | Skin prick test, adjusted (mm) |
|---|---|---|---|---|---|---|---|---|
| 1 | Treatment | 20-24 | Female | White | 44.4 | 0.49 | 109 | 28 |
| 2 | Treatment | 30-34 | Female | White | 4.5 | 0.29 | 40.8 | 10 |
| 3 | Treatment | 20-24 | Female | White | 20.9 | 0.16 | 169 | 5 |
| 4 | Treatment | 15-19 | Female | White | 84.1 | 0.54 | 216 | 10 |
| 5 | Treatment | 5-9 | Male | White | 159 | 0.85 | 338 | 14.5 |
| 6 | Treatment | 10-14 | Male | White | 40.9 | 1.86 | 208 | 7.5 |
| 7 | Treatment | 15-19 | Male | White | 11.2 | 0.16 | 141 | 13 |
| 8 | Treatment | 5-9 | Male | White | 451 | 1.71 | 1524 | 21 |
| 9 | Treatment | 35-39 | Female | Asian | 2.6 | 0.62 | 339 | 13.5 |
| 10 | Placebo | 20-24 | Male | White | 61.4 | 0.09 | 174 | 11 |
| 11 | Placebo | 10-14 | Female | White | 39.1 | 0.18 | 151 | 10 |
| 12 | Placebo | 20-24 | Female | White | 27.3 | 0.37 | 88.1 | 22 |

Supplementary Table 2. Patient clinical outcomes.

| Patient ID | Treatment Group | Cumulative dose consumed at DBFC2 | DBFC2 outcome | Cumulative dose consumed at DBFC3 | DBFC3 outcome | Adverse event count | Therapeutic outcome |
|---|---|---|---|---|---|---|---|
| 1 | Treatment | 4443 | Pass | 4440 | Pass | 605 | Tolerance |
| 2 | Treatment | 4443 | Pass | 4440 | Pass | 269 | Tolerance |
| 3 | Treatment | 4443 | Pass | 4440 | Pass | 61 | Tolerance |
| 4 | Treatment | 4443 | Pass | 4440 | Fail | 306 | Partial tolerance |
| 5 | Treatment | 4440 | Pass | 4440 | Fail | 60 | Partial tolerance |
| 6 | Treatment | 4443 | Pass | 4440 | Fail | 177 | Partial tolerance |
| 7 | Treatment | 943 | Fail | 40 | Fail | 101 | Treatment failure |
| 8 | Treatment | 4443 | Fail | 440 | Fail | 26 | Treatment failure |
| 9 | Treatment | 289.6 | Fail | 1440 | Fail | 497 | Treatment failure |
| 10 | Placebo | 443 | Fail | -- | -- | 81 | Treatment failure |
| 11 | Placebo | 143 | Fail | -- | -- | 42 | Treatment failure |
| 12 | Placebo | 943 | Fail | -- | -- | 25 | Treatment failure |

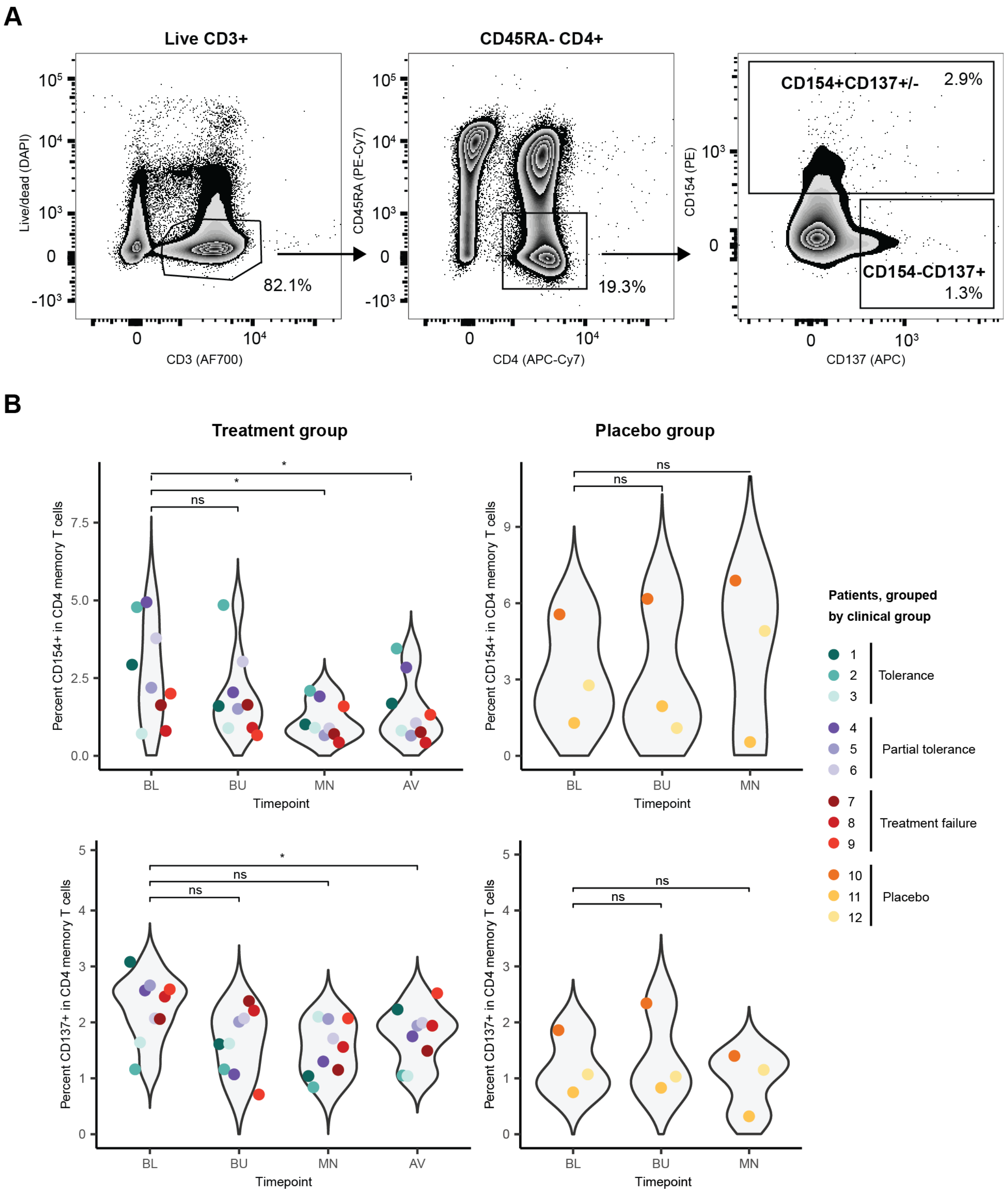


**Supplementary Figure 1. Proportion of CD154+ and CD137+ peanut-reactive T cells. a**, Gating strategy for CD154 and CD137 expression in CD4+ memory T cells from PBMC cultures stimulated with peanut antigen. Data are from a representative patient at baseline. **b**, Percent of CD4+ memory T cells at each time point that are CD154+ (top) or CD137+ (bottom), in patients in the treatment group (left) or placebo group (right). P-values were calculated using a two-sided Wilcoxon rank-sum test and were adjusted with a Bonferroni correction.

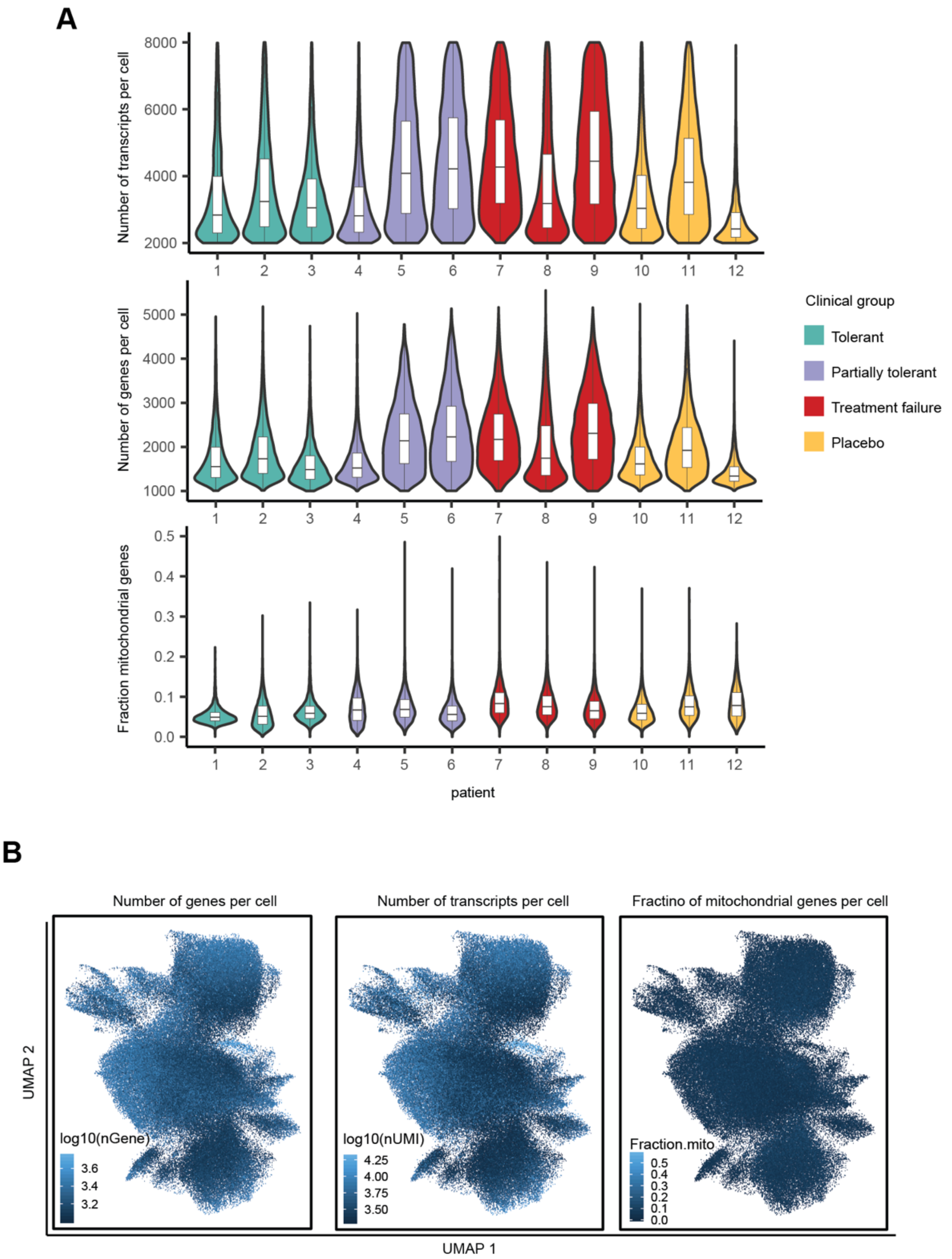


**Supplementary Figure 2. Quality of single-cell RNA-Seq libraries. a**, Quality control metrics of single-cell RNA sequencing libraries: number of UMIs per cell (top), number of genes detected per cell (middle), and fraction of genes detected that are mitochondrial for each cell (bottom). Each violin represents all cells recovered from one patient. **b**, Quality control metrics overlaid on UMAP.

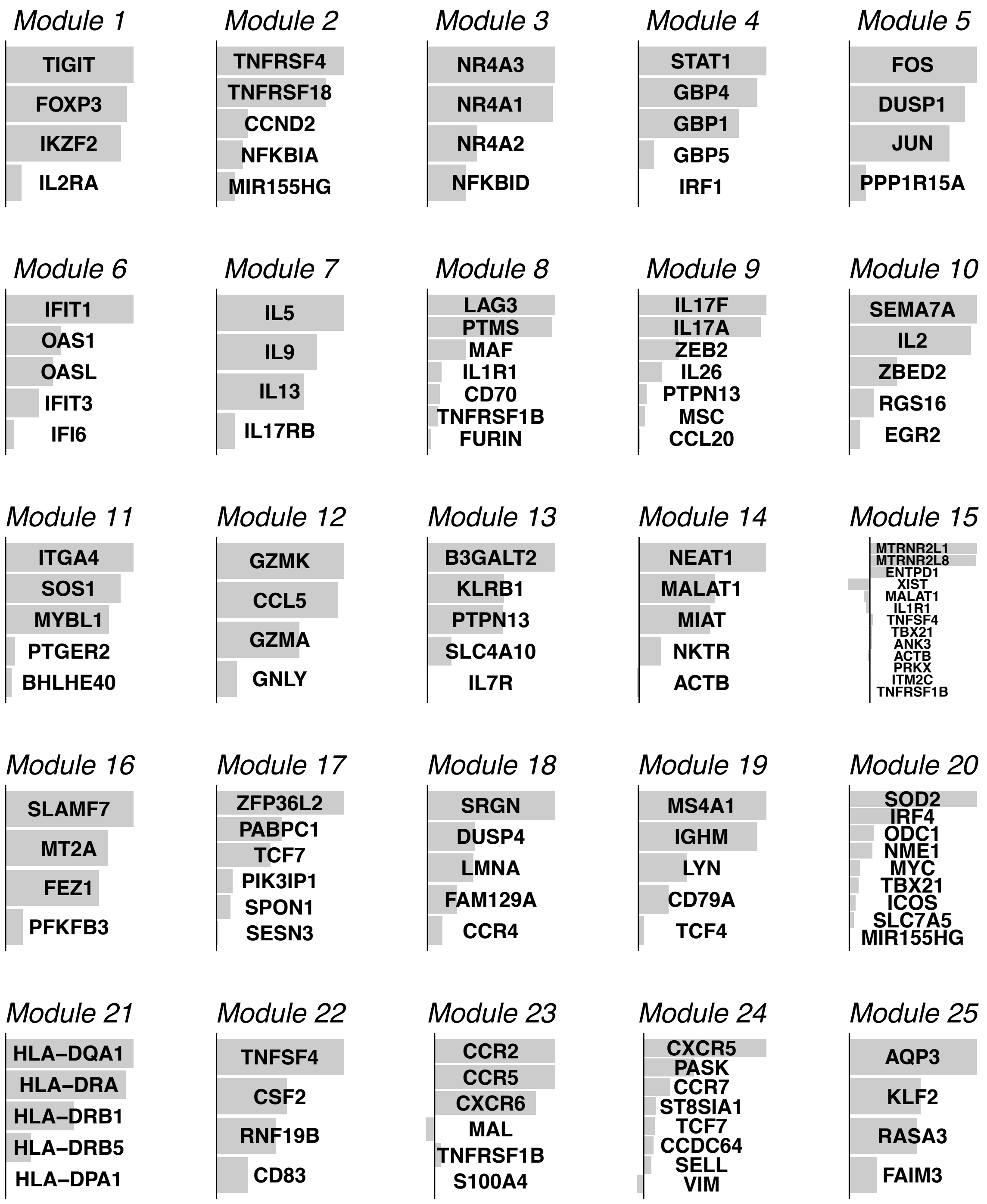


**Supplementary Figure 3. Gene modules 1 through 25.** Gene modules were identified from immune and variable genes in an unsupervised manner using sparse PCA. For each module, the magnitude and direction of each bar represent the weight and sign of each gene in that module, respectively.

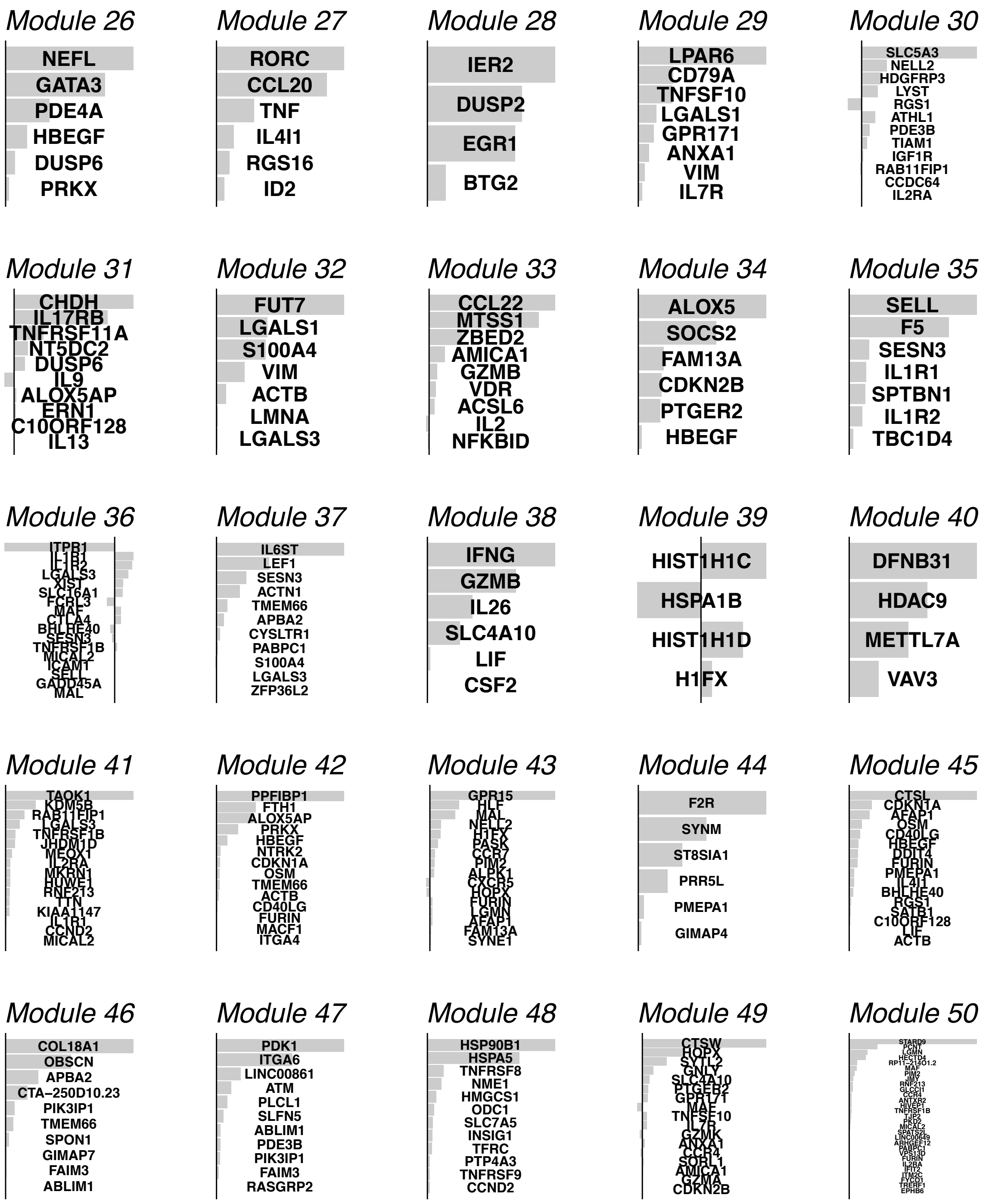


**Supplementary Figure 4. Gene modules 26 through 50.** Gene modules were identified from immune and variable genes in an unsupervised manner using sparse PCA. For each module, the magnitude and direction of each bar represent the weight and sign of each gene in that module, respectively.

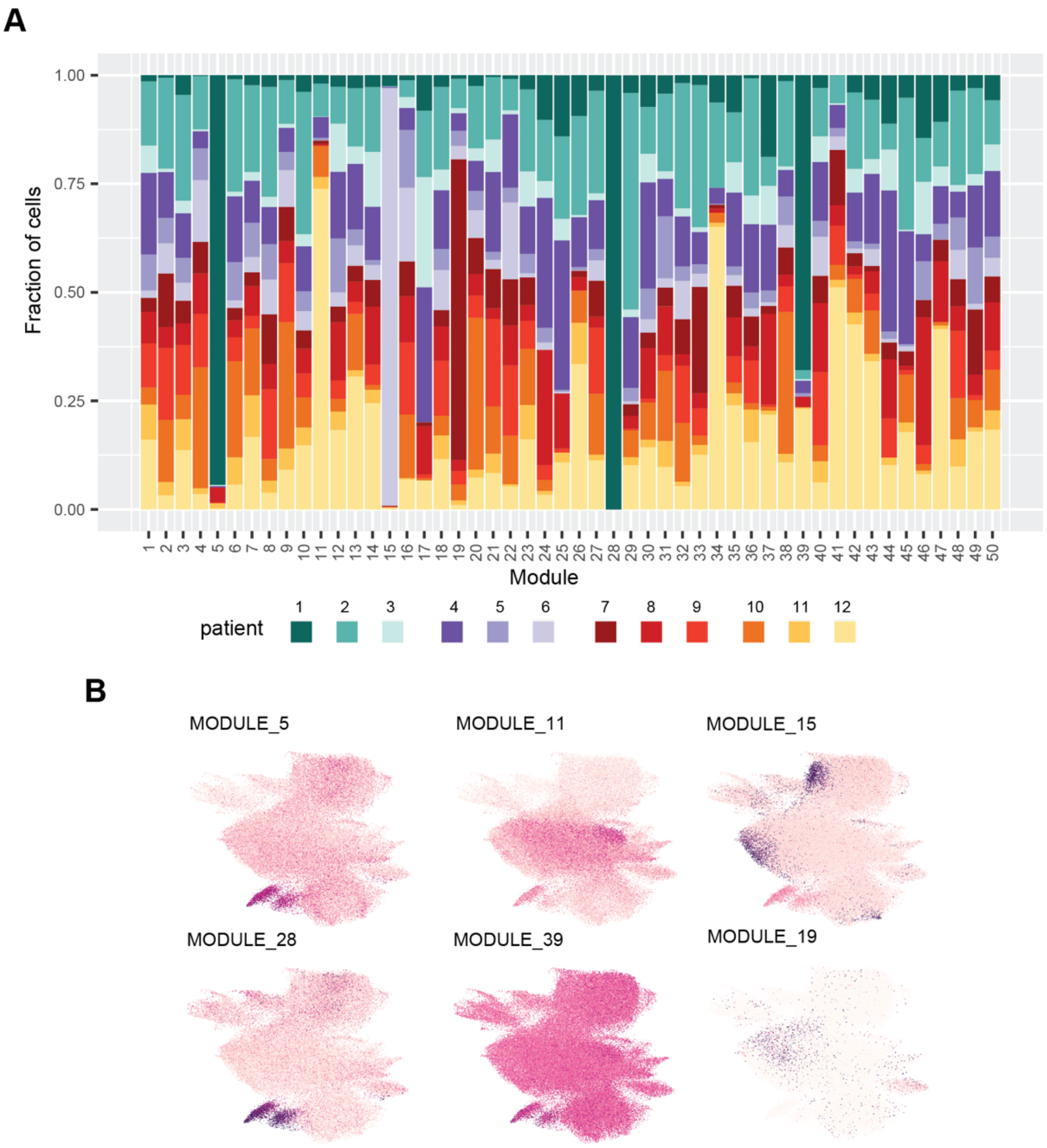


**Supplementary Figure 5. Gene module expression across patients. a**, Distribution of module-expressing cells by patient, for the 50 gene modules. “Module-expressing” cells were determined using the CD154-CD137- cells as described in **Methods**. **b**, UMAP overlay of module expression, and module loadings, for key patient-associated modules.

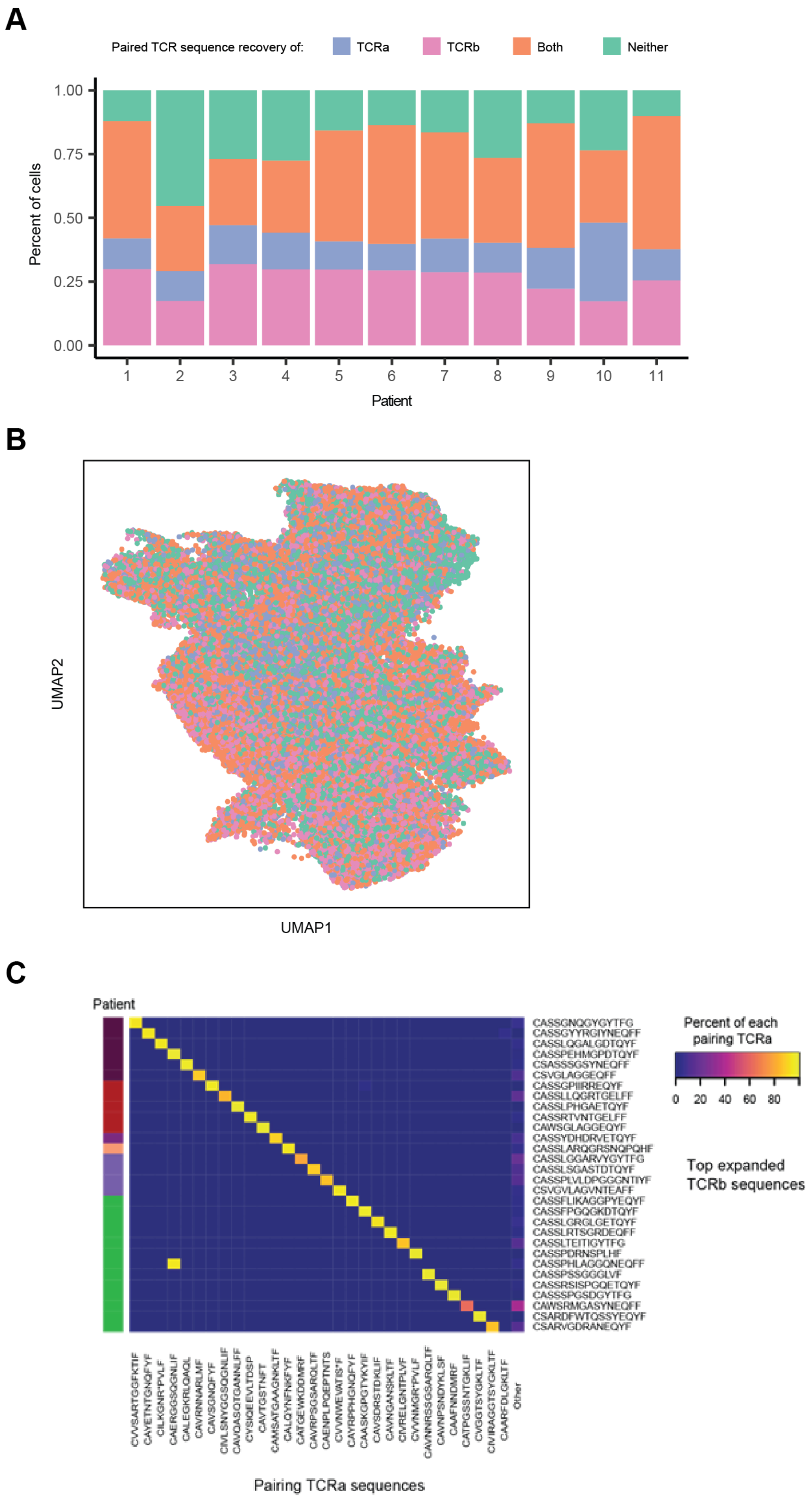


**Supplementary Figure 6. Quality of single-cell TCRα/β sequences. a**, Fraction of cells with each status of TCR sequence recovery for all cells within each patient. **b**, UMAP overlay of TCR recovery status of every cell. **c**, TCRα pairing for the 30 most frequently detected TCRβ sequences. Within cells of each TCRβ sequence, the most frequent pairing TCRα sequences were plotted. The column "Other" captures the fraction of TCRα sequences that were not detected in at least 50% of cells of that TCRβ clonotype. Rows are annotated with the patient in which the TCRβ clonotype was most frequently detected.

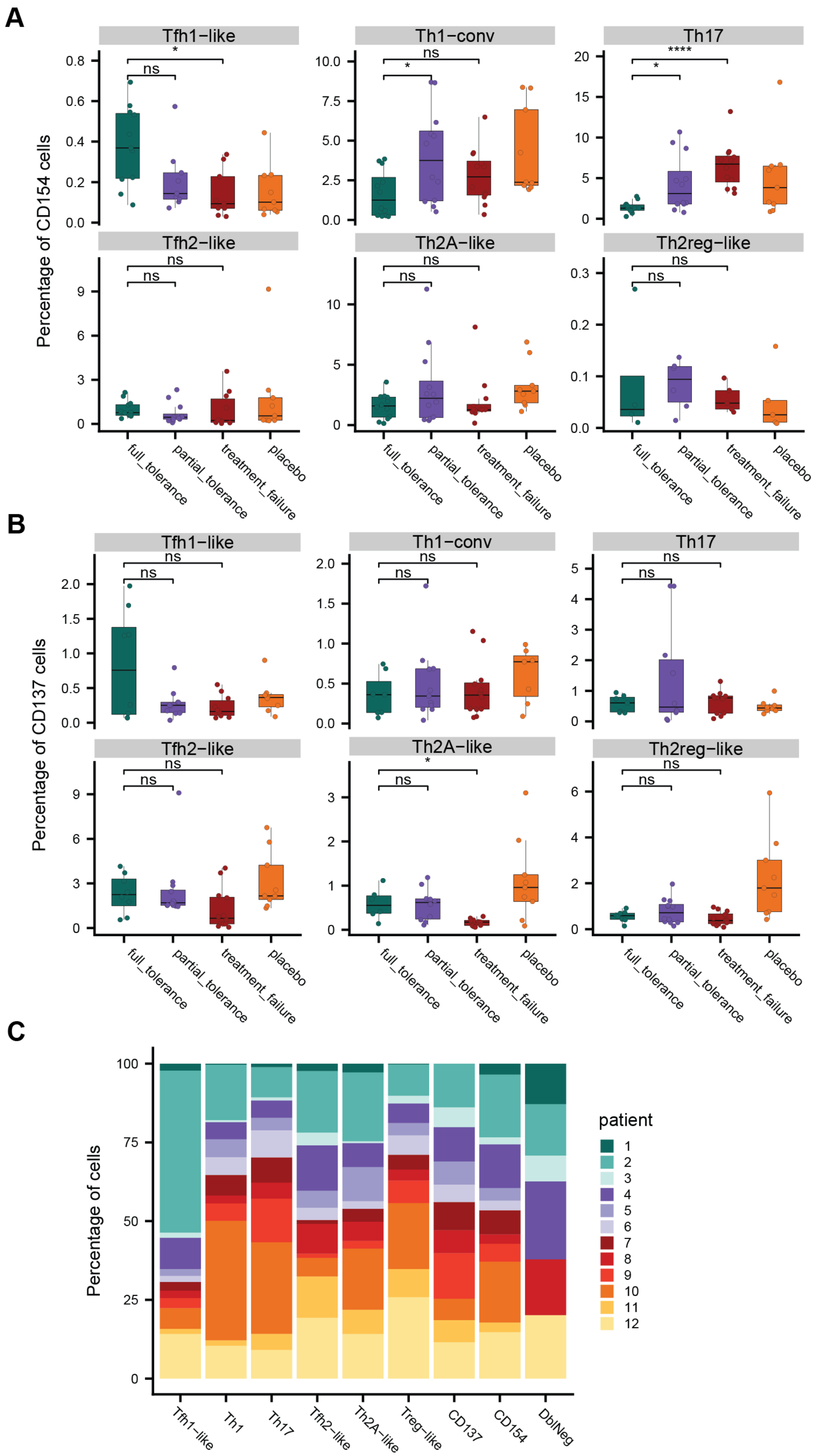


**Supplementary Figure 7. Frequency of T helper subsets by clinical group. a**, Frequency of cells of each T helper subtype among all CD154+ cells for each patient at each timepoint, grouped by clinical group. **b**, Frequency of cells of each T helper subtype among all CD137+ cells for each patient at each timepoint, grouped by clinical group. **c**, Fraction of cells comprising each T helper subtype (and all CD154+, CD137+, or CD154-

CD137- cells, for comparison) from each patient. "*" refers to an adjusted p-value of <0.05 by a Wilcoxon rank-sum test, "**" refers to adjusted p-value of <0.005, and "****" refers to adjusted p-value of <0.0005 (**a**-**b**).

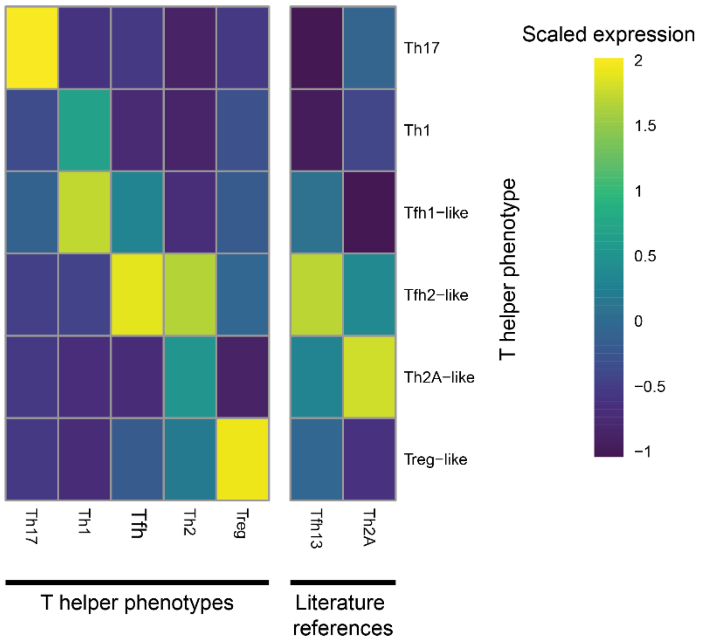


**Supplementary Figure 8. Th1, Th2 and Th17 subsets express previously identified signatures.** Mean expression scores of Th subset signatures identified in previous studies (**Supplementary Table** 4). Th subsets identified in **Figure** 3 (y-axis) were scored for each of the known or previously identified signatures (x-axis) using the AddModuleScore function in Seurat, then averaged within each subset.

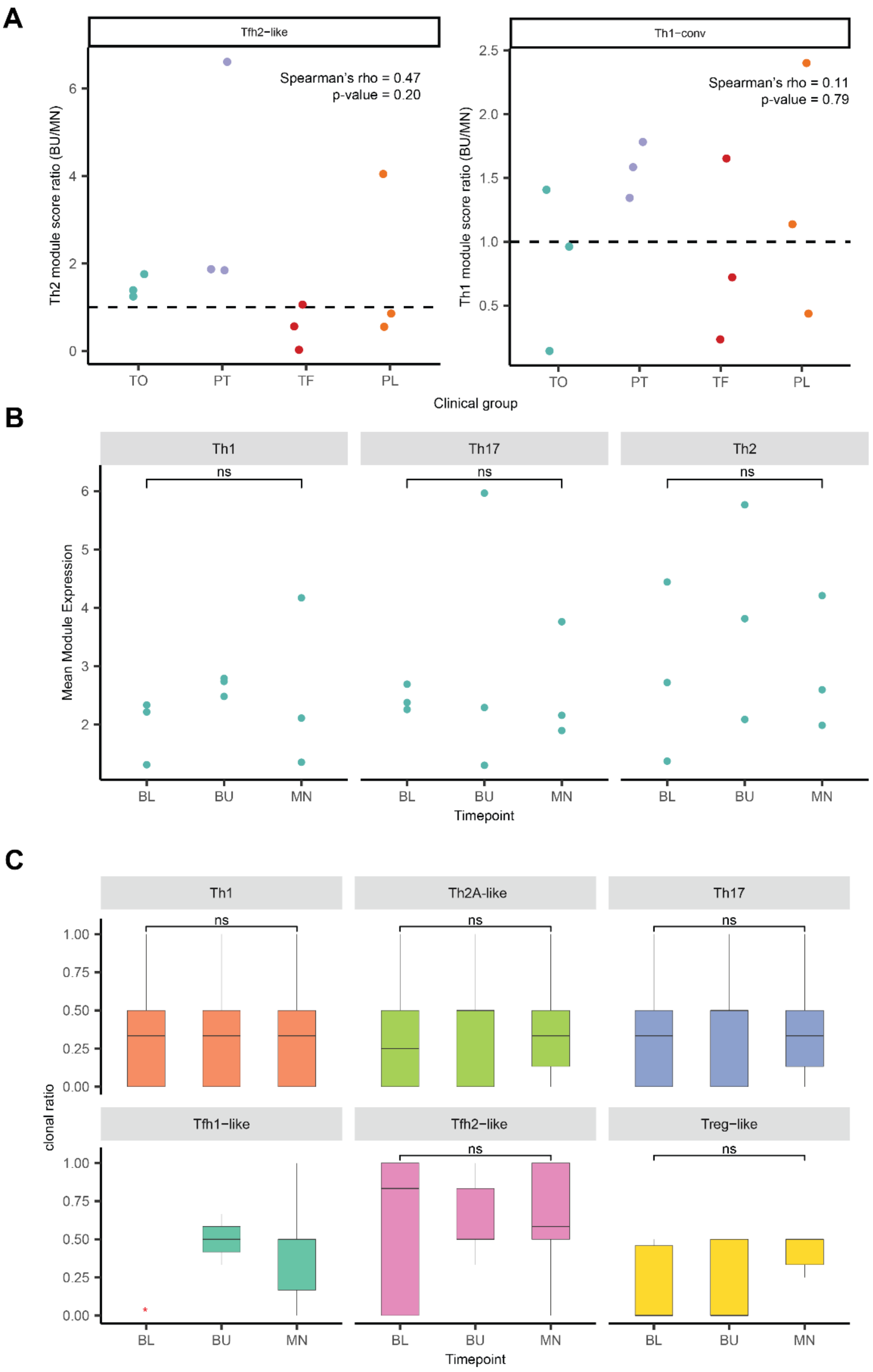


**Supplementary Figure 9. OIT-induced changes by clinical group and in placebo patients. a,** Degree of suppression in Th2A-like cells by clinical group. Ratio of mean Th2 and Th1 module expression in Tfh2-like and Th1-conv clonotypes, respectively, from each patient was calculated between buildup (BU) and maintenance (MN). Spearman’s rho and p-value are from a Spearman correlation test between ratio and outcome within the treatment group (assigning TO as 2, PT as 1, and TF as 0). Dashed line indicates equal module expression between BU and MN (BU/MN ratio of 1). **b**, Mean expression of the Th1, Th2, and Th17 gene modules at baseline (BL), buildup (BU), and maintenance (MN) in CD154+ cells from each of the three placebo patients. **c**, Fractional clonal expression over time of clones from placebo patients in each T helper subset. Fractional clonal expression was defined as the fraction of cells within each clonotype that scored as module-expressing for the relevant gene module (Th2, Th1, or Th17) at a given time point. Clonotypes were only included in the analysis at time points for which they had at least two cells recovered. Red asterisk indicates no clonotypes meeting the criteria were recovered.

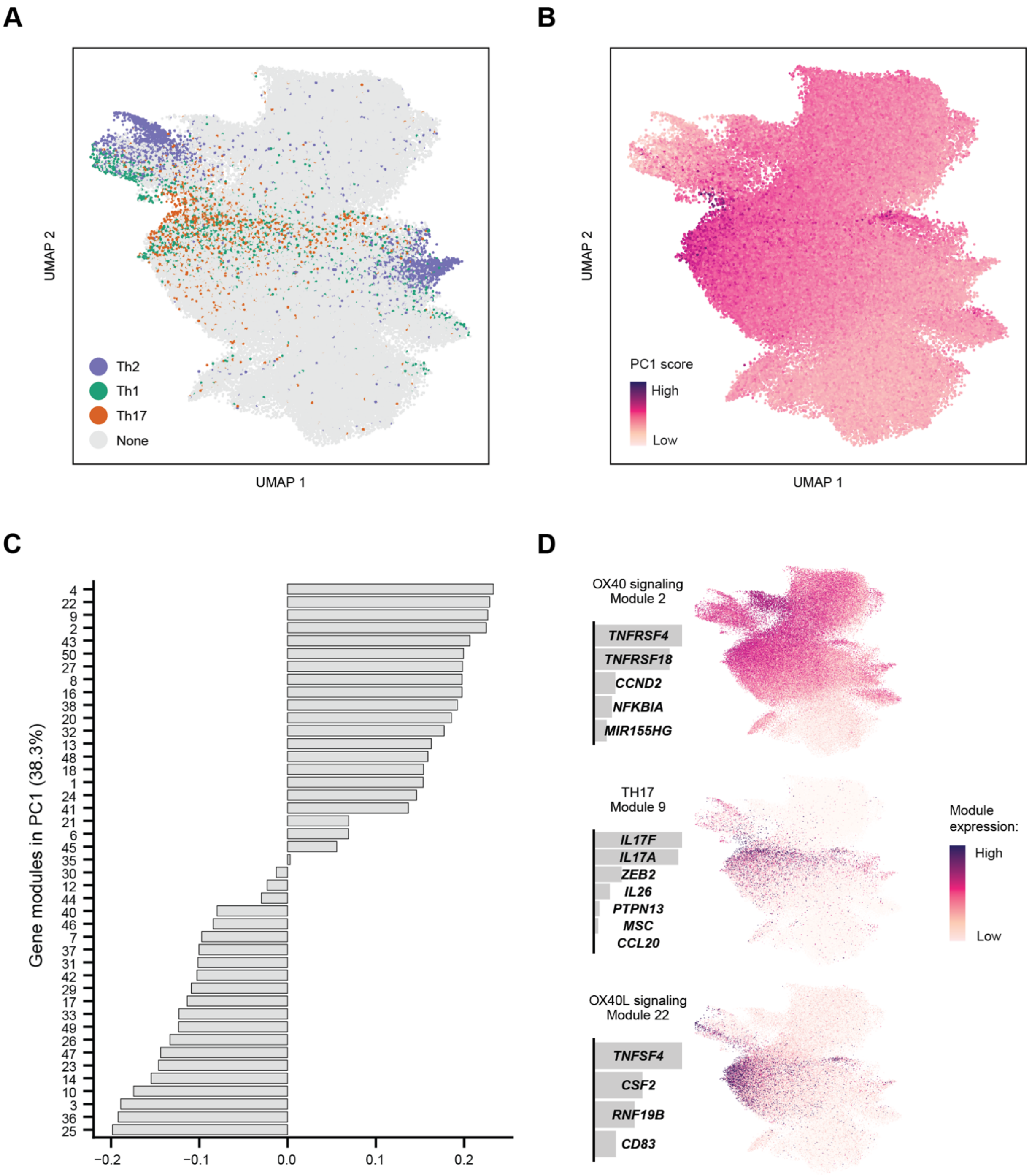


**Supplementary Figure 10. Modules enriched in principal component 1 (PC1) at baseline.** A principal components analysis was done using the 50 gene modules as features and all CD154+ cells at baseline as the input data. **a**, UMAP of all cells colored by Th module classification. Module expression was thresholded as described in **Methods**. **b**, UMAP of all cells colored by PC1 score. **c**, Loadings of each gene module (labeled by number on the y-axis) in principal component 1. **d**, Gene loadings and module expression overlays on the UMAP coordinates, for selected modules positively enriched in PC1.

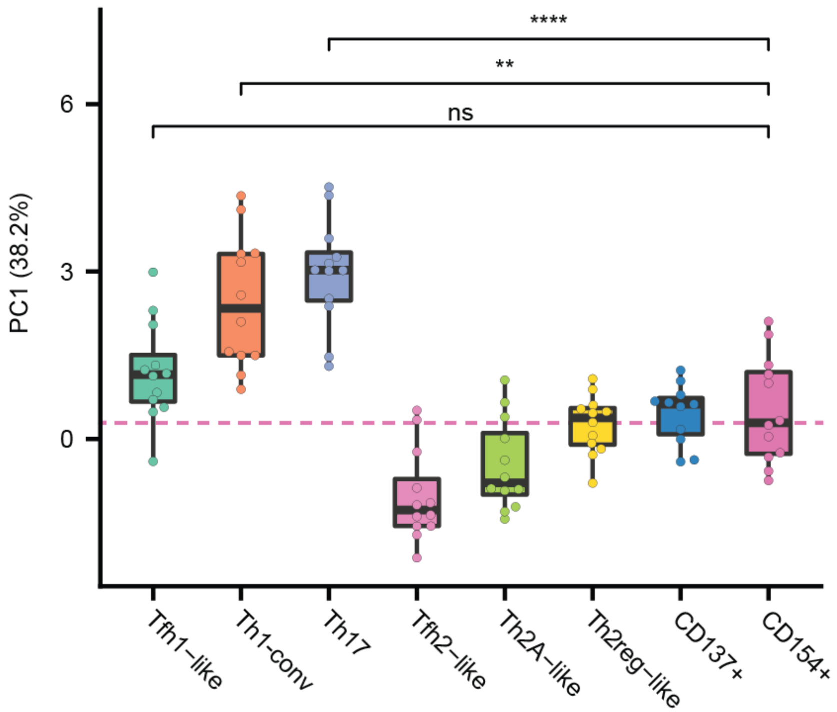


**Supplementary Figure 11. Principal component 1 (PC1) scores of T cell subsets.** A principal components analysis was done using the 50 gene modules as features and all CD154+ cells at baseline as the input data. Each data point represents the mean PC1 score of all cells of a given subset from a single patient at all timepoints. The dotted line represents the median expression of the CD154+ subset. ‘*’ refers to an adjusted p-value of <0.05 by a Wilcoxon rank-sum test, “**” refers to adjusted p-value of <0.005, and “****” refers to adjusted p-value of <0.0005.

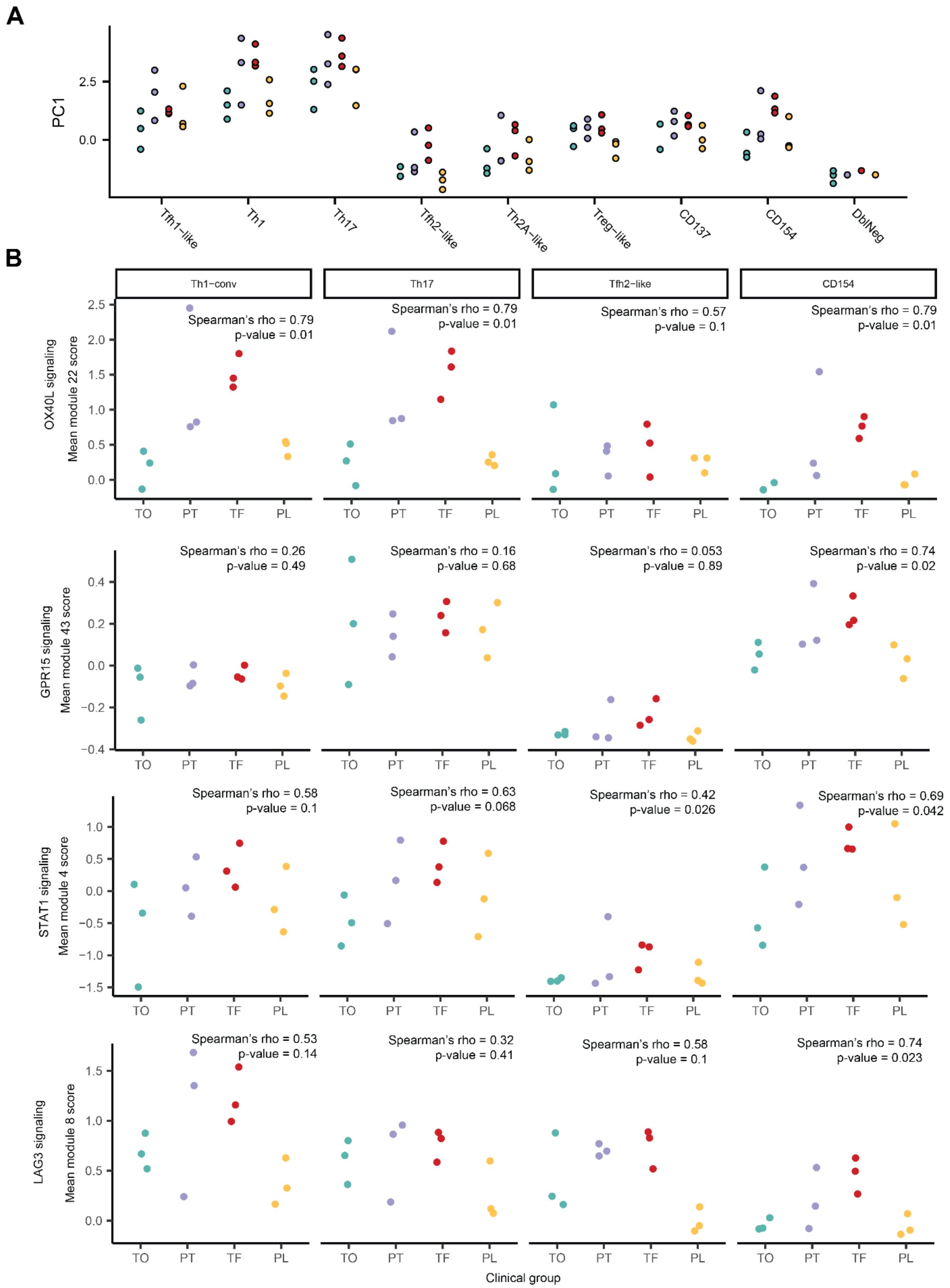


**Supplementary Figure 12. PC1 scores and mean scores of modules enriched in PC1 by cell subset and treatment outcome.** Module expression and PC1 scores of each Th subset shown were averaged by patient at all time points. "CD154" includes all sorted CD154+ cells not categorized as any of the Th subsets shown in **Figure** 3. **a**, PC1 scores, as shown in **Figure** 4D, colored by treatment outcome (green = TO, purple = PT, red = TF, yellow = PL). **b**, Mean module expression of indicated modules in Th1-conv, Th17, Th2A-like, and rest of

CD154+ cells by patient, and colored by outcome. Spearman's rho and p-value are from a Spearman correlation test between mean module scores and treatment outcome, as described in **Methods**.
